# The effect of heat on insecticide-treated net use across Africa: an analysis of 28 malaria-endemic countries from 2011-2022

**DOI:** 10.64898/2026.08.06.26359896

**Authors:** Adrienne Epstein, Max McClure, Paul Krezanoski, Sheri D. Weiser, Isabel Rodriguez-Barraquer

## Abstract

A common barrier to insecticide-treated net (ITN) use, the backbone of malaria prevention, is heat discomfort. Using multilevel regression models, we quantified the association between temperature and ITN use reported in Demographic and Health and Malaria Indicator Surveys from 2011-2022 across 28 countries in sub-Saharan Africa. Marginal predicted probability of use was unimodal with nighttime temperature and decreased above a threshold daytime temperature. For women (n=731,947), temperatures above specified thresholds were associated with lower use, with adjusted odds ratios [aOR] 0.91 for daytime (95% confidence interval [CI] 0.88-0.94) and 0.87 for nighttime (95% CI 0.85-0.91). For children under 5 (n=524,926), corresponding aOR were 0.87 (95% CI 0.84-0.91) and 0.86 (95% CI 0.83-0.90). Considering a counterfactual in which temperatures never exceeded these thresholds, we estimated 25.8 million (95% CI 14.4-38.2 million) cases of malaria attributable to heat-associated non-use. These results demonstrate the importance of considering climate-behavior interactions for malaria control.

## Introduction

Insecticide-treated nets (ITNs) are the most widely used and important tools for malaria prevention, with approximately 73% of households in sub-Saharan Africa owning at least one ITN as of 2023.^2^ Nevertheless, there has been a lack of progress over the last decade in reducing malaria morbidity and mortality.^1^ Because nets are only useful when used regularly, there is reason to believe that non-use may be contributing to at least some apparent ITN failures.^3^ In one study that remotely monitored nightly bednet deployment, ITN non-use exceeded 10% of nights.^4^

In qualitative surveys of ITN use, one of the most commonly cited reasons for non-use is that sleeping under a net feels “too hot,” particularly in warmer months,^5, 6, 7^ ^8, 9^ Although ITNs are unlikely to alter the temperatures experienced by their users, they appear to affect thermal comfort in warm conditions by reducing ventilation and airflow.^10^

As a possible barrier to ITN use, ambient temperature is a compelling, spatiotemporally predictable target for interventions. In some regions it may be addressed through improvements in housing designs or ventilation features,^11^ while in hotter settings, anticipated periods of ITN non-use could inform deployment of alternative or complementary interventions such as spatial emanators. If present, an association between temperature and ITN non-use could also alter predicted associations between climate and malaria transmission risk, as current estimates of the optimal temperature for malaria transmission do not account for human behavior.^12^ This association would have implications for malaria risk maps, seasonal forecasts, and projections under climate change scenarios.

Despite a wealth of qualitative support, the association between population-level ITN use and ambient temperature and the resulting implications for malaria transmission have not to our knowledge been formally evaluated. In this study, we examine the association between remotely sensed daytime and nighttime land surface temperature (LST) and reported ITN use at the subnational level across 28 countries in sub-Saharan Africa.

## Methods

### Data source and participants

This study used data from the nationally representative Demographic and Health Surveys (DHS) and Malaria Indicator Surveys (MIS). The surveys use a stratified two-stage cluster sampling design, first selecting a random sample of enumeration areas (EA), followed by a random sample of households within each EA. All women aged 15 to 49 within selected households are invited to complete a questionnaire. Data derived from surveys include participant age, sex, education status and reported ITN use, household wealth quintile, and region urban/rural status.

We included DHS and MIS conducted in malaria-endemic countries in sub-Saharan Africa from 2011 through 2022. This resulted in 60 surveys from 28 countries (Supplemental Table 1). One eligible country (Namibia) was excluded due to limited temperature variation during survey periods. Analyses included all women aged 15-49 and children under 5 years of age with complete outcome and covariate data.

### Measures

#### Insecticide-treated net use

The primary outcome was a binary variable indicating whether the participant reported sleeping under an ITN the night before the survey. Women aged 15-49 were asked this question for themselves and for all children under five in their household. The primary analysis included all households, regardless of ITN ownership, based on the hypothesis that ownership itself may be influenced by temperature. A sensitivity analysis included only households with at least one ITN.

#### Temperature

The primary exposure was land surface temperature (LST), sourced from Terra Moderate Resolution Imaging Spectroradiometer (MODIS) Land Surface Temperature/Emissivity Daily (MOD11A1)^13^ at five-kilometer resolution. To address missing MODIS observations, we combined Terra and Aqua data within a 30-day window and filled remaining gaps using a 1-pixel neighborhood average applied only to missing locations. We extracted the average monthly daytime and nighttime LST at the EA level for each participant’s survey month. Temperatures were treated as continuous variables and dichotomized into high versus low heat (>30°C for daytime and >20°C for nighttime temperatures). Thresholds were selected based on inflection points identified in generalized additive models (GAMs), described below. As a sensitivity analysis, we assessed alternative cut points defining three categories of nighttime temperatures. A further sensitivity analysis used a cut point of 35°C for daytime temperatures, established from previous “extreme heat” definitions.^14^ Because daytime and nighttime temperatures may influence behavior through distinct mechanisms, both were entered simultaneously into all models.

#### Covariates

We adjusted for factors that may influence ITN use and be associated with living in warmer environments. Because areas above a certain temperature may have lower malaria risk due to vector and parasite suitability constraints,^12^ and perceived malaria risk is a known predictor of net use,^15^ we adjusted all models for malaria prevalence at the EA level using Malaria Atlas Project (MAP) estimates of *Plasmodium falciparum* prevalence in 2-10 year-olds (categorized as <10%, 10 to <20%, 20 to <30%, and ≥30%).^16^ All models also adjusted for survey month (to account for seasonality), household urban/rural status, wealth quintile,^17^ and EA-level elevation.^18^ As a sensitivity analysis, we adjusted for cluster-level malaria prevalence measured via rapid diagnostic tests or microscopy within the DHS/MIS surveys (rather than via the MAP). This measure was only available in a subset of surveys, and the DHS/MIS sampling design is not powered to produce precise cluster-level prevalence estimates, but we included this analysis to directly capture local transmission intensity and seasonality beyond what the MAP modeled surfaces provide.

Separate models were fit for women and children to test for differences in association. Models including women aged 15-49 additionally adjusted for participant age (5-year categories) and education level (none, primary, secondary, higher). Models including children under five adjusted for child sex, age (in months), and caregiver age (5-year categories). Pooled models including women and children adjusted for sex and age (continuous).

### Statistical Analysis

#### Temperature-ITN associations

We used multilevel logistic regression models fit separately to the women and children datasets to estimate the association between temperature and ITN use. We fit models both pooled across countries and stratified by country. Pooled models included survey-level fixed effects to account for country- and time-specific differences; stratified models included survey-round fixed effects within each country. All models used robust standard errors clustered at the EA level.

To model potential nonlinearities in the associations between ITN use and temperature, we used GAMs with logit links. To flexibly account for nonlinear temperature effects, we included separate cubic-regression-spline smooths for daytime LST and nighttime LST. Additional models assessed the joint effects of daytime and nighttime temperature via a two-dimensional smooth for their interaction.

To assess the associations between binary categorizations of temperature (based on the thresholds defined above) and ITN use, we used multilevel logistic regression models. Models produced odds ratios (ORs) and 95% confidence intervals (CIs) for each temperature category. We also derived marginal predicted probabilities of ITN use by temperature category. Effect modification was assessed by including interaction terms between high daytime and nighttime temperatures. Interactions were evaluated on the multiplicative scale using joint p-values for the interaction terms.

Daytime and nighttime LST were moderately correlated (r = 0.45); variance inflation factors (VIFs) indicated no collinearity concerns in the pooled model. In country-stratified models, the daytime-by-nighttime interaction inflated VIFs and was excluded, retaining only main effects.

#### Estimating cases attributable to heat-associated ITN non-use

We quantified malaria cases from 2011-2022 attributable to reduced ITN use associated with high temperatures. For each country, we fit logistic regression models of ITN use combining data from children under 5 and women aged 15-49 with binary indicators of daytime and nighttime temperatures above their thresholds (30°C and 20°C, respectively). From each model, we extracted the coefficient vector and variance-covariance matrix. We used these coefficients to reconstruct predicted probabilities of ITN use for three strata: (1) neither daytime nor nighttime above threshold, (2) daytime above threshold, and (3) nighttime above threshold.

We combined these ITN use probabilities with spatially and temporally resolved temperature and malaria incidence data to estimate excess cases attributable to heat-related reductions in ITN use. For each grid cell and year, we used monthly MODIS land surface temperature rasters to calculate the proportion of months with daytime temperatures exceeding 30°C and nighttime temperatures exceeding 20°C. We obtained annual *Plasmodium falciparum* incidence count surfaces from the Malaria Atlas Project.^16^

The malaria cases attributable to ITN non-use were estimated as the difference between observed incidence and the incidence assuming the dichotomized temperature thresholds were not exceeded. For each grid cell, the observed population-level ITN use was modeled as a weighted average of stratum-specific ITN use probabilities, weighted by the local fraction of time in each temperature category. The counterfactual scenario assumed that temperatures never exceeded thresholds, such that ITN use equaled the below-threshold probability. We estimated counterfactual malaria incidence by rescaling observed incidence according to the ratio of protective coverage under the two scenarios, using a protective efficacy of 0.45, derived from the rate ratio of 0.55 (95% CI: 0.48-0.64) for uncomplicated *P. falciparum* malaria episodes reported in a Cochrane review of randomized trials of ITNs versus no nets.^19^ Specifically, for a grid cell with observed incidence , fraction of time above threshold , ITN use probability above threshold , and below threshold , cases not averted were calculated as:

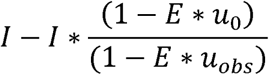

where *u_obs_* = (1-*f*) * *u*_0_ + *f* * *u*_1_ and *E* = 0.45. Under this model, full ITN coverage would reduce malaria incidence to 55% of the level expected without nets, consistent with the Cochrane estimate. The excess represents the additional malaria cases arising because high temperatures reduce ITN use below what it would be in the absence of extreme heat. Daytime and nighttime contributions were computed from the same model using a single set of simulation draws to preserve the covariance structure of the shared intercept and then summed. Grid-cell-level estimates were aggregated to country-year totals.

To quantify uncertainty, we used a Monte Carlo simulation approach. For each country, we drew 1,000 coefficient vectors from the multivariate normal distribution defined by the estimated coefficients and their variance-covariance matrix. Each draw was transformed through the logistic function to obtain simulated ITN use probabilities, which were then propagated through the cases-not-averted formula. The 2.5th and 97.5th percentiles of the resulting distribution of total cases not averted defined the 95% confidence intervals. Finally, for each country we calculated the proportion of *P. falciparum* cases attributable to heat-related reductions in ITN use by dividing cases not averted by the total case count from the corresponding years in the World Malaria Report.^2^

## Results

### Sample characteristics

The analytic sample included 731,947 women aged 15-49 and 524,926 children from 28 countries (see Supplemental Table 1 sample sizes by survey). Table 1 shows individual-, household- and enumeration area-level characteristics of the sample. 47.1% of women and 52.7% of children reported using an ITN the night prior to the survey (for these estimates by country, see Supplemental Table 2).

**Table 1.** Descriptive characteristics of women 15-49 and children under 5 included in the analysis.

|  |  |  |
| --- | --- | --- |
|  | Women<br>(n=731,947) | Children<br>(n=524,926) |
| **Individual-level characteristics** |  |  |
| Age in years, mean (SD) | 28.5 (9.5) | 2.0 (1.4) |
| Male, % (n) |  | 50.4% (264,502) |
| Education level, % (n) |  |  |
| None | 31.4% (230,160) |  |
| Primary | 33.6% (246,235) |  |
| Secondary | 30.2% (221,205) |  |
| Higher | 4.7% (34,347) |  |
| Used insecticide-treated net in previous night, % (n) | 47.1% (345,037) | 52.7% (276,427) |
| **Household- level characteristics (n = 529,925 households)** |  |  |
| Urban, % (n) | 35.1% (185,738) |  |
| Wealth quintile, % (n) |  |  |
| Poorest | 21.6% (114,640) |  |
| Poorer | 19.6% (103,845) |  |
| Middle | 19.3% (102,209) |  |
| Richer | 19.5% (103,485) |  |
| Richest | 20.0% (105,746) |  |
| Daytime temperature during survey month >30°C, % (n) | 51.0% (270,308) |  |
| Daytime temperature during survey month in °C, mean (SD) [range] | 31.2 (5.7) [12.1-52.0] |  |
| Nighttime temperature during survey month >20°C, % (n) | 38.8% (205,744) |  |
| Nighttime temperature during survey month in °C, mean (SD) [range] | 18.5 (3.7) [3.6-29.3] |  |
| Precipitation during survey month in millimeters, mean (SD) | 108.1 (129.3) |  |
| Household elevation in meters, mean (SD) | 691.6 (607.7) |  |
| *P falciparum* prevalence among 2–10-year-olds, mean (SD) | 0.18 (0.14) |  |

Household-level mean daytime and nighttime temperatures during the survey month were 31.2°C (standard deviation [SD] 5.7) and 18.5°C (SD 3.7) respectively. See Supplemental Figure 1 for temperature ranges by country. As described in the methods, daytime temperatures during survey month were classified as “hot” (defined as an average monthly daytime LST > 30°C) in 51.0% of households containing both women and children. Nighttime temperatures during survey month were hot (defined as an average monthly nighttime LST > 20°C) in 38.8% of these households. Mean *P. falciparum* prevalence among 2-10-year-olds was 0.18 (SD 0.14) in enumeration areas containing both women and children.

### Temperature and ITN use

We first used generalized additive models (GAMs) to assess nonlinear relationships between temperature and the marginal predicted probability of ITN use. We found the association between daytime temperature and ITN use declined with increasing daytime LST above approximately 30°C for both women and children. At lower temperatures, the association with ITN use either did not change or slightly increased with increasing temperature. Only a small proportion of observations fell below the curves’ respective maxima, limiting power to estimate this association (7% for women and 2% for children). The relationship between ITN use and nighttime LST was unimodal, peaking near 20°C for both women and children (Fig. 1). Results from country-level GAMs are shown in Supplemental Figures 2 and 3. Allowing for nonlinear interactions between daytime and nighttime temperatures demonstrated a decreasing probability of ITN use with increasing daytime temperatures when nighttime temperatures lay between approximately 15°C and 27°C and a unimodal relationship between nighttime temperatures and the probability of ITN use with a relative maximum near 17°C independent of daytime temperature (Fig. 2).

**Figure 1.**
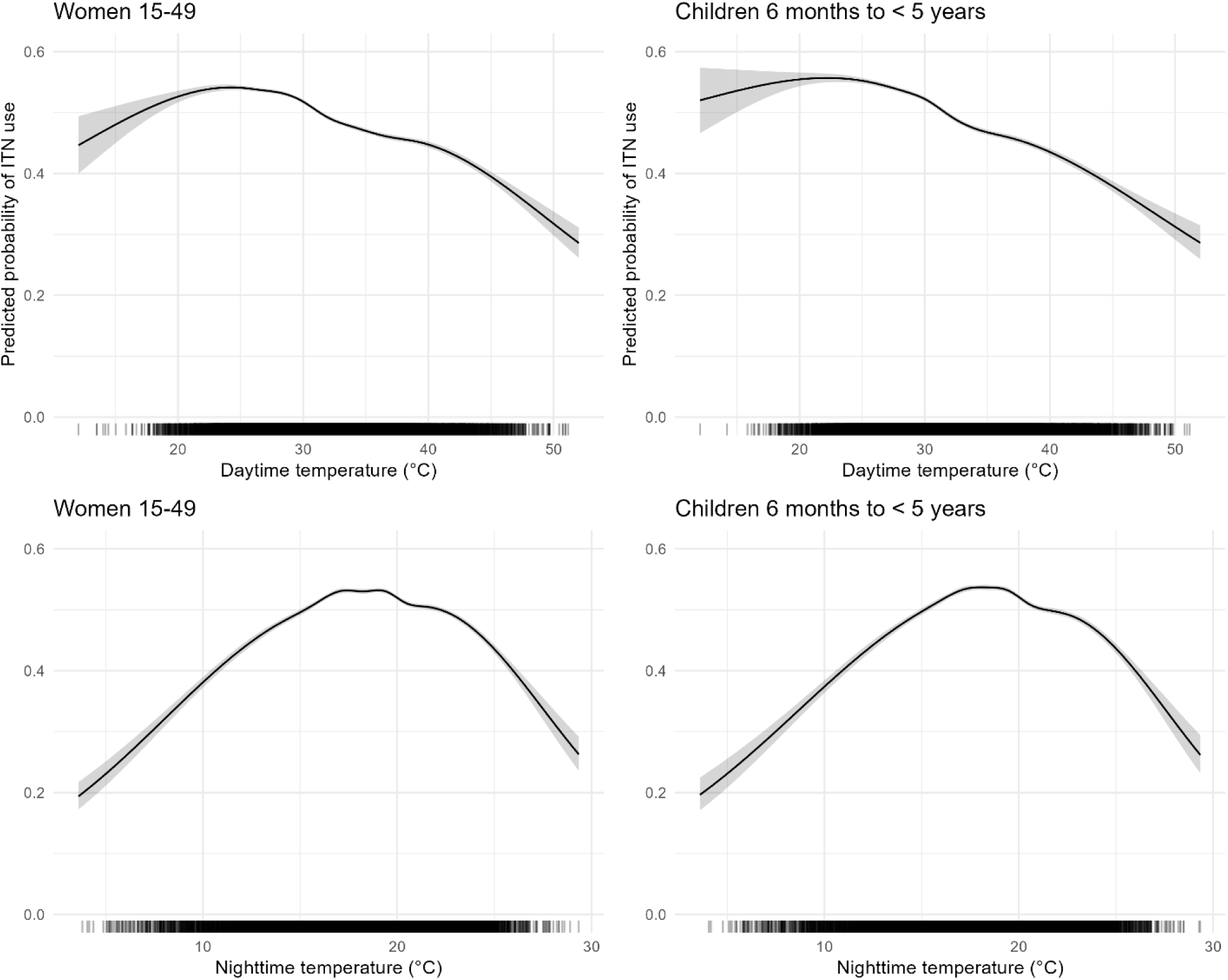
Predicted ITN use as a function of daytime and nighttime temperature for women aged 15-49 and children under five. Curves represent generalized additive model estimates with 95% confidence intervals.

**Figure 2.**
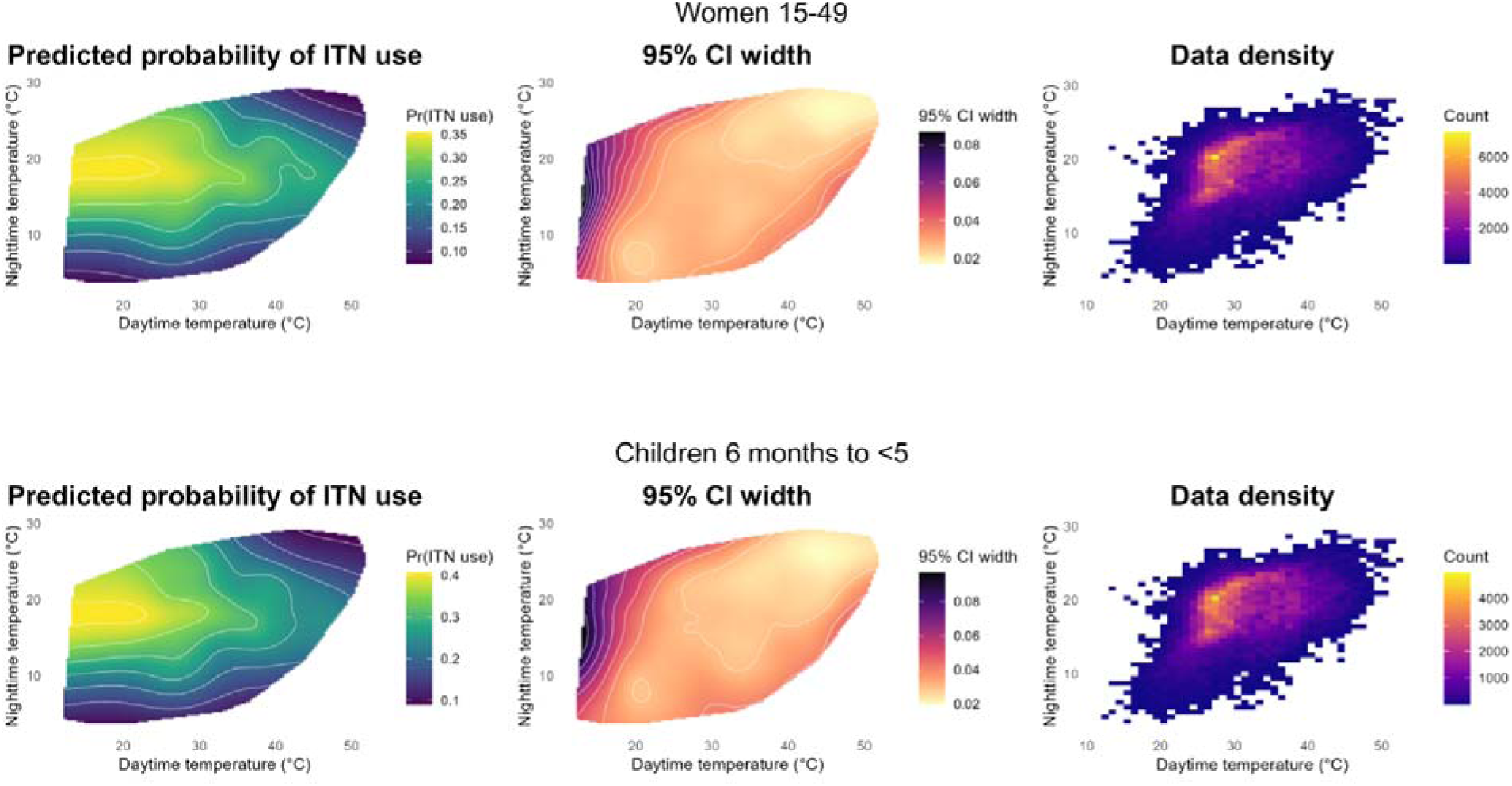
Predicted probabilities of ITN use, 95% confidence-interval widths, and observed temperature distributions for women aged 15-49 and children under five, estimated using generalized additive models. Predictions are shown only over the range of observed daytime and nighttime temperatures.

**Figure 3.**
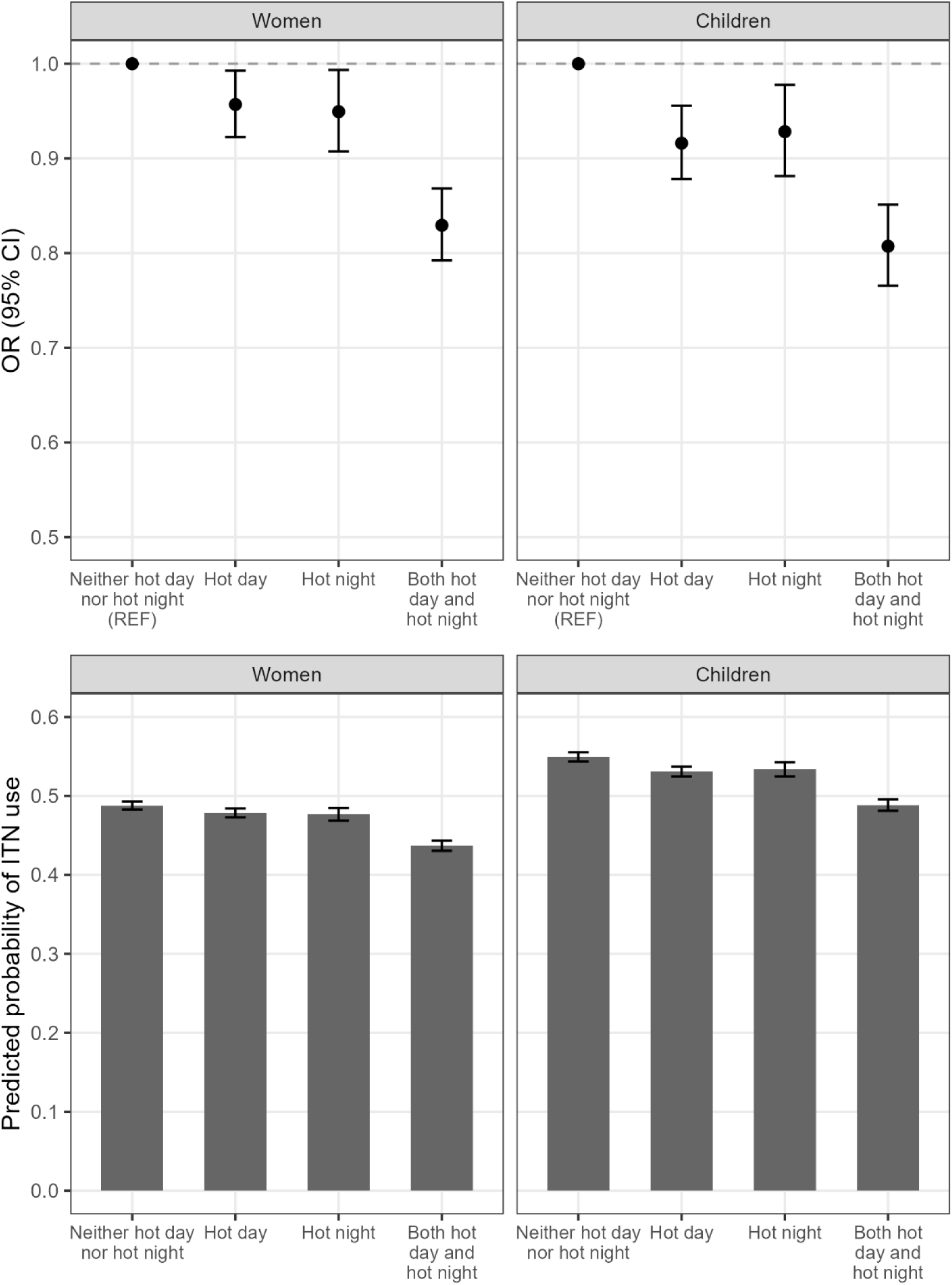
Odds ratios and predicted probabilities of ITN use demonstrating the interaction between daytime and nighttime temperature.

We used multilevel logistic regression models to evaluate associations between “hot” days or nights (as defined above) and odds of survey-reported ITN use (Table 2), adjusting for relevant confounders. Both associations were independently significant for both women and children. For women, we found that “hot” daytime and nighttime temperatures were associated with a 9% (95% CI 6-12%) and 13% (95% CI 9-15%) decrease in the odds of using a net, respectively. For children, the corresponding decreases were 13% (95% CI 9-16%) and 14% (95% CI 10-17%).

**Table 2.** Associations between heat and insecticide-treated net use among women and children.

|  |  |  |
| --- | --- | --- |
| Population | Unadjusted | Adjusted |
|  | Odds ratio (95% CI) |  |
| Women aged 15-49 years (n=731,947) |  |  |
| Daytime temp >30°C | 0.86*** (0.84, 0.89) | 0.91*** (0.88, 0.94) |
| Nighttime temp >20°C | 0.97* (0.94, 0.99) | 0.87*** (0.85, 0.91) |
| Children under 5 years of age (n=524,926) |  |  |
| Daytime temp >30°C | 0.85*** (0.82, 0.88) | 0.87*** (0.84, 0.91) |
| Nighttime temp >20°C | 0.95** (0.92, 0.99) | 0.86*** (0.83, 0.90) |
| Coefficients are presented as odds ratios with 95% confidence intervals in parentheses. Models include adjustment for survey, survey month, wealth quintile, urban, precipitation in the survey month, parasite prevalence, and elevation at the enumeration area level. Models for women include additional adjustment for education level and age category. Models for children include additional adjustment for child age, child sex, and caregiver age category. Standard errors are clustered at the enumeration area level. Asterisks denote level of significance ***p<0.001 **p<0.01 *p<0.05 |  |  |

As a sensitivity analysis, we examined whether the association between nighttime temperature and ITN use differed across three categories: cold nights (<15°C), moderate nights (15-20°C, reference), and hot nights (>20°C). This specification was motivated by the GAM results suggesting that ITN use peaked at moderate temperatures and declined at both extremes. In the three-category analysis, both cold nights (<15°C) and hot nights (>20°C) were associated with lower ITN use relative to moderate nights (15-20°C), consistent with the unimodal pattern observed in the GAM (Supplemental Table 3). Notably, observations with cold nighttime temperatures (<15°C) were concentrated in a small number of countries with high-altitude regions, including Kenya, Madagascar, Rwanda, and Burundi.

We assessed for an interaction between daytime and nighttime LST by evaluating multiplicative interactions between the two temperatures. For both groups, the combination of hot day and night resulted in lower odds of ITN use than a hot day or night alone, compared to neither hot day nor hot night: for women, the combination was associated with a 16% decrease in the odds of using a net (95% CI 12-20%, interaction p<0.001), and for children a 19% decrease (95% CI 15-23%, interaction p<0.001) (Fig. 3).

Considering that transmission intensity may be a significant confounder of the association between temperature and net use, we repeated the analysis replacing MAP-modeled prevalence with cluster-level malaria prevalence measured directly within DHS/MIS surveys (Supplemental Figure 4). Using the full subsample with DHS/MIS prevalence data, results were nearly identical to the main analysis using MAP prevalence. We further restricted to clusters with increasingly large sample sizes (≥2, ≥10, ≥20, ≥30, and ≥40 tested individuals) to improve the precision of cluster-level prevalence estimates. The daytime temperature OR remained stable near 0.85-0.90 across all thresholds and losing precision only at the most restrictive cutoffs. The nighttime temperature association strengthened as cluster size increased, with the OR declining from ∼0.85 in the full sample to ∼0.50 at cluster sizes ≥30 and ≥40. Further sensitivity analyses restricted to households that owned at least one ITN (Supplemental Table 4) and using 35°C as the daytime cut point (Supplemental Table 5) showed consistent results, both in directionality and relative magnitude.

Country-level analyses showed broadly consistent evidence that extreme heat was associated with reduced ITN use (Fig. 4). Hot daytime or nighttime temperatures alone were generally associated with modest declines, while concurrent daytime and nighttime heat produced the largest marginal risk differences in the majority of countries with available data (ranging from approximately −5 to −25 percent). Country-exposure combinations omitted from Fig. 4 were not estimable due to limited within-country variation in temperatures. Statistically significant associations in the opposite direction, where increasing temperature led to increases in ITN use, were seen in Gabon (hot days only), Burundi and Ghana (both hot nights only).

**Figure 4.**
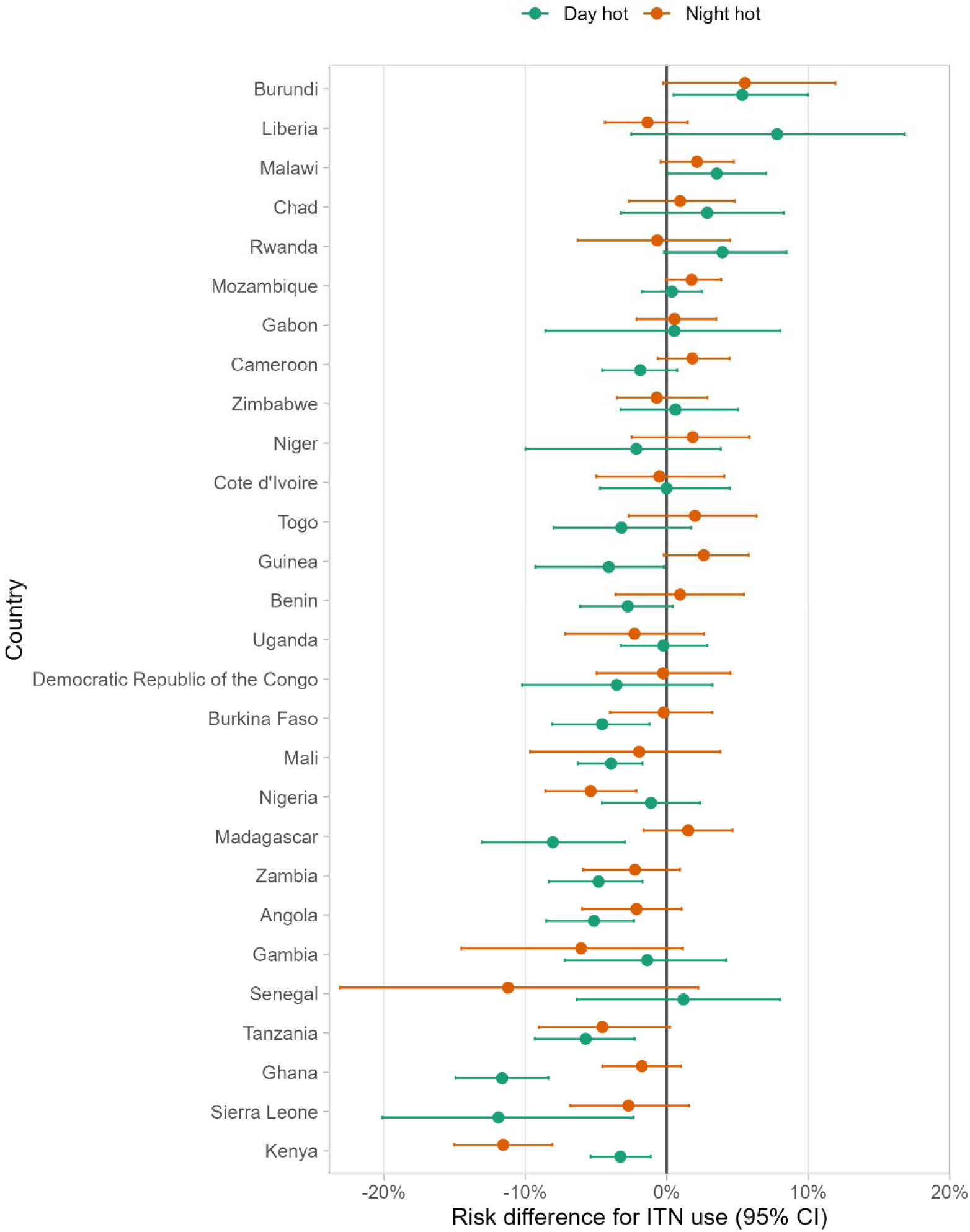
Country-specific risk differences (95% CI) for the associations of daytime and nighttime heat with ITN use among women aged 15-49 and children under five.

### Heat-associated ITN non-use and malaria cases not averted

Using the outputs from these models, we estimated overall and country-level malaria cases attributable to heat-associated ITN non-use from 2011-2022. Across the 28 countries analyzed, we estimated a total of 25.8 million attributable cases not averted (95% CI 14.4-38.2 million), accounting for 1.06% (95% CI 0.59-1.57%) of malaria cases during that period. Most countries showed a positive burden, although the 95% confidence intervals for cases not averted excluded zero only in Angola, Burkina Faso, Ghana, Kenya, Madagascar, Nigeria, Sierra Leone, Tanzania and Zambia. In Burundi, Cameroon, Chad, Gabon, Malawi, Mozambique, Rwanda and Zimbabwe the estimated effect was negative, indicating fewer cases due to higher estimated ITN use under heat conditions, with 95% confidence intervals excluding zero only in Burundi and Malawi. Total case contribution estimates ranged from an increase of 14.0 million cases in Nigeria to a decrease of 559,000 cases in Chad. The percent of national cases attributable to high temperatures ranged from 2.85% in Mali to −1.40% in Chad (Supplemental Table 6, Fig. 5). Results by country and year are shown in Supplemental Table 7.

**Figure 5.**
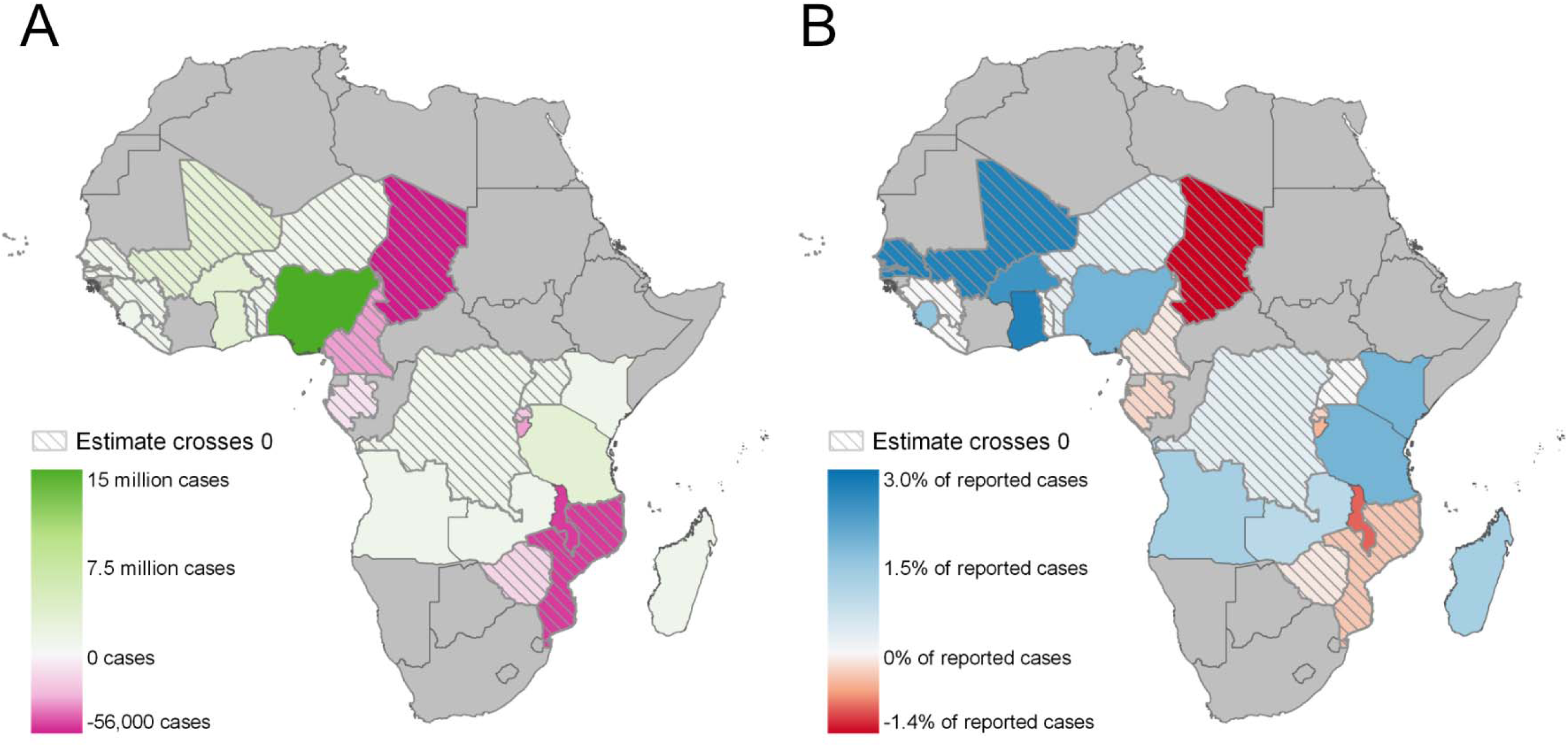
Country-level malaria cases not averted attributable to heat-related reductions in ITN use, 2011-2022, expressed as (A) absolute cases not averted and (B) cases not averted expressed as a percentage of reported malaria cases from the 2024 World Malaria Report. Hatched countries have estimates with confidence intervals crossing zero.

## Discussion

In this study, we found that the association between temperature and ITN use was nonlinear, with daytime temperatures above 30°C and nighttime temperatures above 20°C associated with a decrease in reported ITN use in women of childbearing age and children under five years across a wide geographic range. The combination of hotter daytime and nighttime temperatures resulted in a lower probability of ITN use than either alone. Based on these associations, we estimate that ITN non-use associated with high temperatures resulted in 25.8 million malaria cases across 28 countries in sub-Saharan Africa from 2011-2022. To our knowledge, this is the first large-scale quantitative evaluation of the influence of temperature on ITN use and its likely effect on disease burden.

While our analysis does not address the mechanisms by which high temperatures might affect ITN use, qualitative studies frequently cite discomfort due to high temperatures as one of the most common reasons for ITN non- or under-use.^5, 6, 7^ Social, economic and psychological stressors attributable to climate change and associated extreme weather events may also undermine health-related behaviors more generally, potentially including the use of ITNs.^20^ It is possible that lower perceived or actual malaria risk during hot, dry seasons may play a role in explaining the observed pattern,^5, 7^ though we adjusted for regional malaria prevalence and season. Notably, this approach would not necessarily address the role of nuisance biting by non-malaria vector species, which also affects ITN use^6^ and would also be expected to vary with climate.

The apparent multiplicative interaction between the effects of hot daytime and nighttime temperatures may reflect the cumulative effect of thermal stress over a 24-hour period or may suggest the combination of daytime and nighttime temperatures is more representative of the conditions present when the average individual is preparing to use an ITN. Interactions between the effects of daytime and nighttime temperatures have also been reported for other heat-related disease outcomes,^21^ potentially attributable to the loss of a nighttime recovery period. The loss of this relationship between daytime temperatures and ITN use when nighttime temperatures were very hot or very cold (< 15°C and > 27°C) suggests such extreme conditions may outweigh daytime temperature effects, although our power was limited over these ranges.

Contrary to the negative relationship between increasing temperatures and ITN use at moderate to high temperatures, at nighttime temperatures below approximately 17-18°C we found that the marginal predicted probability of ITN use decreased with decreasing temperature. This pattern may in part reflect the low risk of malaria and/or nuisance biting in cold climates, particularly in deserts and high-elevation regions with cold nighttime temperatures, as suggested by the decrease in effect size after adjusting for a set of covariates that included elevation and malaria prevalence.

The negative association between higher temperatures and ITN use held for most, but not all, countries. Excess malaria cases were attributable to heat-associated ITN non-use in 20 of 28 countries; in the remaining eight (Burundi, Cameroon, Chad, Gabon, Malawi, Mozambique, Rwanda and Zimbabwe), higher temperatures were instead associated with greater ITN use and thus fewer attributable cases, though confidence intervals excluded zero only in Burundi and Malawi.

Inspection of country-level smooths (Supplemental Figs 2-3) indicates that most of these reversals reflect the limitations of our binary temperature threshold rather than genuinely protective behavior. In Malawi, Mozambique, Zimbabwe and Cameroon, daytime ITN use was unimodal but peaked above our 30°C threshold. Malawi’s nighttime smooth was similarly peaked. A monotonic increase in ITN use was confined to nighttime temperatures and reached significance only in Burundi. Its mechanism is unclear, though Burundi and Rwanda are the only highland countries in this group, which may distinguish them from the lowland-dominated pooled estimate; both countries reported few temperatures above the nighttime “hot” thresholds (4.9% and 0.4%, respectively). The unexpected results in Chad and Gabon instead reflect data limitations: Chad recorded few daytime temperatures below the 30°C threshold (3.9%), while Gabon’s estimates were imprecise and near-null. Qualitative studies of ITN non-use in Burundi, Malawi and Rwanda suggest that, as in other regions, ITNs are perceived as uncomfortable in hot weather.^9, 22, 23^

Our findings have several implications for malaria control measures. Associations between temperature and ITN use may inform the timing of interventions and the interpretation of malaria trials that span multiple seasons or climate regimes. In regions where high indoor temperatures are unavoidable, it may be reasonable to prioritize novel vector control measures such as spatial emanators to mitigate the expected decrease in LLIN use. Elsewhere, lowering nighttime temperatures inside sleeping structures may improve ITN use. A variety of housing modifications have been proposed that improve ventilation and cool indoor temperatures for this purpose, including screened windows, elevated structures and permeable walls.^11, 25^

It is unclear, however, whether such measures would translate to overall lower malaria risk. The relationship between temperature and transmission suitability of *P. falciparum* by *Anopheles gambiae*, the principal malaria vector in sub-Saharan Africa, is thought to be unimodal.^26^ Extreme indoor heat due to metal roofing, which our analysis suggests would be associated with a lower probability of ITN use, has also been associated with lower vector survival.^27^ Models accounting for empirically derived temperature-dependent vector and parasite life traits suggest that the optimal temperature for malaria transmission is as low as 25°C, meaning that measures designed to improve ITN use through cooling could in some cases paradoxically improve predicted vectorial capacity. To this point, DHS and MIS surveys are conducted during peak malaria season for each study region, which already provides evidence that the occasional extreme temperatures we describe were not sufficient to mitigate vector exposure.

The geographical breadth of this analysis is one of its strengths, but the necessarily coarse spatial scales at which we evaluate land surface temperature and ITN use mean that the associations we describe are ecological. As noted above, we are unable to comment on the route by which temperature leads to decreased ITN use, and it is possible that spatially or temporally fine-scale variations in malaria risk drive ITN use that we cannot evaluate at the EA level. Possible effect modifiers that are beyond the scope of this study include ITN type and the existence of housing modifications likely to affect interior temperatures. Our analysis also relies on self-reported ITN use, which has been shown to overestimate ITN use relative to objective evaluations.^29^

Finally, our estimate of malaria cases not averted due to high temperatures is a simplification. In particular, we assume a constant reduction in the rate of *P. falciparum* cases with ITN use across all countries, when there is likely significant local variation in intervention effectiveness driven partly by differences in endemicity levels and ITN types.^30^ Furthermore, the heat-ITN associations used to estimate cases not averted were derived from women aged 15-49 and children under 5 and extrapolated to the full population. However, this inference is unlikely to substantially bias our conclusions as household sleeping arrangements and behaviors are likely correlated within families^31^ and children under 5 represent the subgroup most influential for the calculation of malaria burden.

In summary, we find support for an association between higher ambient temperatures and decreased ITN use across much of sub-Saharan Africa. ITN non-use associated with higher temperatures may contribute significantly to malaria burden in endemic countries. Further research is needed to incorporate these findings into the implementation of targeted malaria control measures and current climate-informed models of malaria transmission.

## Data Availability

All data used in the present study are available from the Demographic and Health Surveys (https://www.dhsprogram.com/).

