## Supplements for "The effect of heat on insecticide-treated net use across Africa: an analysis of 28 malaria-endemic countries from 2011-2022"

| **Supplemental Table 1.** List of surveys and sample sizes included in analysis. | | |
| --- | --- | --- |
| **Survey** | **N women aged 15-49** | **N children under 5** |
| Angola DHS 2015-16 | 13028 | 12596 |
| Angola MIS 2011 | 7984 | 7194 |
| Benin DHS 2017-18 | 14901 | 11913 |
| Burkina Faso DHS 2021 | 17123 | 11500 |
| Burkina Faso MIS 2014 | 7846 | 6287 |
| Burkina Faso MIS 2017-18 | 6918 | 5328 |
| Burundi DHS 2016-17 | 16967 | 12231 |
| Burundi MIS 2012 | 5108 | 4011 |
| Cameroon DHS 2011 | 14314 | 10044 |
| Cameroon DHS 2018 | 11376 | 7846 |
| Cameroon MIS 2022 | 4988 | 3474 |
| Chad DHS 2014-2015 | 17660 | 16772 |
| Democratic Republic of the Congo DHS 2013-2014 | 17128 | 15585 |
| Cote d'Ivoire DHS 2011-2012 | 9702 | 6800 |
| Gabon DHS 2012 | 7748 | 5263 |
| Gabon DHS 2019-21 | 8534 | 5458 |
| Gambia DHS 2019-2020 | 11620 | 7746 |
| Ghana DHS 2014 | 7674 | 4566 |
| Ghana DHS 2022 | 14882 | 8966 |
| Ghana MIS 2016 | 4877 | 2920 |
| Ghana MIS 2019 | 4823 | 2687 |
| Guinea DHS 2012 | 8254 | 5778 |
| Guinea DHS 2018 | 10552 | 7091 |
| Guinea MIS 2021 | 5203 | 3529 |
| Kenya DHS 2014 | 30713 | 19787 |
| Kenya DHS 2022 | 31973 | 18725 |
| Kenya MIS 2015 | 5375 | 3472 |
| Kenya MIS 2020 | 6770 | 3562 |
| Liberia DHS 2013 | 8185 | 6487 |
| Liberia DHS 2019-2020 | 7662 | 4995 |
| Liberia MIS 2011 | 3253 | 2651 |
| Liberia MIS 2016 | 3783 | 2569 |
| Liberia MIS 2022 | 3670 | 2322 |
| Madagascar DHS 2021 | 18755 | 11679 |
| Madagascar MIS 2011 | 8057 | 6012 |
| Madagascar MIS 2013 | 7954 | 5305 |
| Madagascar MIS 2016 | 10649 | 6873 |
| Malawi DHS 2015-16 | 24526 | 16361 |
| Malawi MIS 2012 | 2903 | 2205 |
| Malawi MIS 2014 | 2896 | 2045 |
| Malawi MIS 2017 | 3807 | 2280 |
| Mali DHS 2012-2013 | 10387 | 9502 |
| Mali DHS 2018 | 9784 | 8691 |
| Mali MIS 2015 | 7755 | 7365 |
| Mali MIS 2021 | 10568 | 9202 |
| Mozambique DHS 2011 | 13652 | 10193 |
| Mozambique DHS 2022-23 | 13023 | 8724 |
| Mozambique MIS 2018 | 6085 | 4382 |
| Niger DHS 2012 | 11117 | 11528 |
| Niger MIS 2021 | 5795 | 4795 |
| Nigeria DHS 2013 | 37283 | 27461 |
| Nigeria MIS 2015 | 6733 | 5318 |
| Rwanda DHS 2014-15 | 13468 | 7513 |
| Rwanda DHS 2019-2020 | 14620 | 7749 |
| Senegal MIS 2020-21 | 10159 | 6160 |
| Sierra Leone DHS 2013 | 13206 | 8455 |
| Sierra Leone DHS 2019 | 10118 | 5926 |
| Tanzania DHS 2015-16 | 13051 | 9569 |
| Tanzania MIS 2017 | 9873 | 7310 |
| Togo DHS 2013-14 | 9403 | 6446 |
| Togo MIS 2017 | 4624 | 3263 |
| Uganda DHS 2016 | 18178 | 14349 |
| Uganda MIS 2014-15 | 5245 | 4452 |
| Uganda MIS 2018-19 | 8167 | 6225 |
| Zambia DHS 2013-14 | 16202 | 12477 |
| Zambia DHS 2018 | 13392 | 9201 |
| Zimbabwe DHS 2015 | 9918 | 5755 |
| **TOTAL** | 731947 | 524926 |

| **Supplemental table 2.** Percent of respondents who used an ITN the prior night, by country. | | |
| --- | --- | --- |
| **Country** | **Women aged 15-49** | **Children under 5** |
| Angola | 23.5 | 28.9 |
| Benin | 75.4 | 80.1 |
| Burkina Faso | 67.3 | 73.5 |
| Burundi | 42.2 | 47.9 |
| Cameroon | 34.7 | 40.2 |
| Chad | 21.2 | 22.2 |
| Democratic Republic of the Congo | 55.6 | 61.1 |
| Cote d'Ivoire | 39.6 | 47.3 |
| Gabon | 26.9 | 34.4 |
| Gambia | 45.7 | 52.5 |
| Ghana | 44.7 | 56.1 |
| Guinea | 29.4 | 34.3 |
| Kenya | 48 | 54 |
| Liberia | 45.1 | 49.7 |
| Madagascar | 61 | 67.1 |
| Malawi | 47.7 | 53.7 |
| Mali | 72.7 | 78.8 |
| Mozambique | 43.3 | 47.8 |
| Niger | 40.5 | 41.4 |
| Nigeria | 34.1 | 39.9 |
| Rwanda | 59 | 66.8 |
| Senegal | 53.7 | 56 |
| Sierra Leone | 53.1 | 61.6 |
| Tanzania | 54.4 | 57.8 |
| Togo | 47.6 | 56.1 |
| Uganda | 65.3 | 71 |
| Zambia | 48.1 | 53 |
| Zimbabwe | 10.4 | 11.9 |

**Supplemental Table 3.** Associations between nighttime temperature (3-categories) and insecticide-treated net use among women and children combined (n=1,248,919)

|  | Unadjusted | Adjusted |
| --- | --- | --- |
|  | Odds ratio (95% CI) | |
| Nighttime temp <15°C | 0.65*** (0.62, 0.67) | 0.74*** (0.71, 0.77) |
| Nighttime temp 15°C to 20°C | REF | REF |
| Nighttime temp >20°C | 0.91*** (0.88, 0.94) | 0.86*** (0.83, 0.89) |

| **Supplemental table 4.** Associations between heat and insecticide-treated net use among women and children, restricting to households that own at least one ITN. | | |
| --- | --- | --- |
| Population | Unadjusted | Adjusted |
|  | Odds ratio (95% CI) | |
| Women aged 15-49 years (n= 501,805) | | |
| Daytime temp >30°C | 0.87*** (0.85, 0.90) | 0.88*** (0.85, 0.91) |
| Nighttime temp >20°C | 0.97* (0.94, 0.99) | 0.93*** (0.90, 0.97) |
| Children under 5 years of age (n=371,639) | | |
| Daytime temp >30°C | 0.86*** (0.83, 0.89) | 0.85*** (0.82, 0.89) |
| Nighttime temp >20°C | 0.95** (0.91, 0.98) | 0.93** (0.88, 0.97) |
| Coefficients are presented as odds ratios with 95% confidence intervals in parentheses.  Models include adjustment for survey, survey month, wealth quintile, urban, precipitation in the survey month, parasite prevalence, and elevation at the enumeration area level.  Models for women include additional adjustment for education level and age category. Models for children include additional adjustment for child age, child sex, and caregiver age category.  Standard errors are clustered at the enumeration area level.  Asterisks denote level of significance ***p<0.001 **p<0.01 *p<0.05 | | |

| **Supplemental table 5.** Associations between heat and insecticide-treated net use among women and children, with 35°C as the cut point for daytime temperatures. | | |
| --- | --- | --- |
| Population | Unadjusted | Adjusted |
|  | Odds ratio (95% CI) | |
| Women aged 15-49 years (n=731,947) | | |
| Daytime temp >35°C | 0.82*** (0.80, 0.84) | 0.86*** (0.83, 0.90) |
| Nighttime temp >20°C | 0.97* (0.94, 0.99) | 0.87*** (0.84, 0.90) |
| Children under 5 years of age (n=524,926) | | |
| Daytime temp >35°C | 0.78*** (0.75, 0.81) | 0.82*** (0.78, 0.85) |
| Nighttime temp >20°C | 0.96* (0.93, 0.99) | 0.87*** (0.83, 0.90) |
| Coefficients are presented as odds ratios with 95% confidence intervals in parentheses.  Models include adjustment for survey, survey month, wealth quintile, urban, precipitation in the survey month, parasite prevalence, and elevation at the enumeration area level.  Models for women include additional adjustment for education level and age category. Models for children include additional adjustment for child age, child sex, and caregiver age category.  Standard errors are clustered at the enumeration area level.  Asterisks denote level of significance ***p<0.001 **p<0.01 *p<0.05 | | |

| Supplemental Table 6. Estimated cases not averted due to high temperatures from 2011-2022, by country, expressed in absolute terms and as percents relatives to cases from the same time period reported in the 2024 World Malaria Report. | | | | | |
| --- | --- | --- | --- | --- | --- |
| Country | Cases not averted due to high temperatures 2011-2022 | Lower bound of cases not averted | Upper bound of cases not averted | Total number of cases from 2011-2022 | Percent of cases due to high temperatures 2011-2022 (95% CI) |
| Angola | 808,506 | 337,153 | 1,466,181 | 67,606,658 | 1.20% (0.50%, 2.17%) |
| Benin | 178,361 | -1,115,009 | 1,409,652 | 54,415,369 | 0.33% (-2.05%, 2.59%) |
| Burkina Faso | 2,323,912 | 226,982 | 4,501,718 | 96,054,551 | 2.42% (0.24%, 4.69%) |
| Burundi | -157,587 | -270,978 | -59,372 | 31,555,201 | -0.50% (-0.86%, -0.19%) |
| Cameroon | -61,053 | -504,966 | 371,816 | 83,667,904 | -0.07% (-0.60%, 0.44%) |
| Chad | -559,204 | -1,568,791 | 566,347 | 39,930,511 | -1.40% (-3.93%, 1.42%) |
| Democratic Republic of the Congo | 1,131,597 | -2,415,326 | 4,304,711 | 319,135,832 | 0.35% (-0.76%, 1.35%) |
| Gabon | -7,574 | -43,734 | 28,946 | 4,411,738 | -0.17% (-0.99%, 0.66%) |
| Gambia | 131,117 | -26,281 | 297,305 | 5,531,026 | 2.37% (-0.48%, 5.38%) |
| Ghana | 2,293,248 | 1,339,962 | 3,217,756 | 83,834,165 | 2.74% (1.60%, 3.84%) |
| Guinea | 78,545 | -312,422 | 569,259 | 50,419,094 | 0.16% (-0.62%, 1.13%) |
| Cote d’Ivoire | 127,558 | -1,006,771 | 1,253,355 | 89,297,094 | 0.14% (-1.13%, 1.40%) |
| Kenya | 741,709 | 538,186 | 928,426 | 38,488,417 | 1.93% (1.40%, 2.41%) |
| Liberia | 28,998 | -129,860 | 193,591 | 17,998,243 | 0.16% (-0.72%, 1.08%) |
| Madagascar | 354,769 | 63,646 | 629,161 | 31,289,832 | 1.13% (0.20%, 2.01%) |
| Malawi | -472,786 | -844,812 | -124,474 | 50,214,794 | -0.94% (-1.68%, -0.25%) |
| Mali | 2,019,292 | -47,762 | 4,306,634 | 70,753,347 | 2.85% (-0.07%, 6.09%) |
| Mozambique | -477,124 | -1,085,656 | 127,778 | 114,674,176 | -0.42% (-0.95%, 0.11%) |
| Niger | 320,338 | -1,790,918 | 3,154,926 | 83,207,361 | 0.38% (-2.15%, 3.79%) |
| Nigeria | 14,032,985 | 3,937,349 | 24,763,154 | 738,839,616 | 1.90% (0.53%, 3.35%) |
| Rwanda | -21,831 | -44,223 | 1,030 | 9,043,536 | -0.24% (-0.49%, 0.01%) |
| Senegal | 394,076 | -327,949 | 1,076,498 | 14,668,573 | 2.69% (-2.24%, 7.34%) |
| Sierra Leone | 454,057 | 53,784 | 845,417 | 31,394,233 | 1.45% (0.17%, 2.69%) |
| Togo | 14,710 | -604,491 | 711,377 | 29,287,771 | 0.05% (-2.06%, 2.43%) |
| Uganda | 200,105 | -492,057 | 888,054 | 138,212,640 | 0.14% (-0.36%, 0.64%) |
| Tanzania | 1,584,905 | 837,077 | 2,290,866 | 82,447,408 | 1.92% (1.02%, 2.78%) |
| Zambia | 378,879 | 144,296 | 653,892 | 40,835,220 | 0.93% (0.35%, 1.60%) |
| Zimbabwe | -11,837 | -172,309 | 143,272 | 18,988,996 | -0.06% (-0.91%, 0.75%) |
| TOTAL | 25,828,670 | 14,358,107 | 38,181,405 | 2,436,203,303 | 1.06% (0.59%, 1.57%) |

| **Supplemental Table 7**. Estimated cases not averted due to high temperatures from 2011-2022, by country and year, expressed in absolute terms and as percents relatives to cases from the same time period reported in the 2024 World Malaria Report. | | | | | | | | |
| --- | --- | --- | --- | --- | --- | --- | --- | --- |
| **Country** | **Year** | **Cases due to non-use** | **Cases due to non-use (lower 95% CI)** | **Cases due to non-use (Upper 95% CI)** | **Cases reported in WMR** | **Proportion of cases due to non-ITN use** | **Proportion of cases due to non-ITN use (lower 95% CI)** | **Proportion of cases due to non-ITN use (upper 95% CI)** |
| Angola | 2011 | 28014.89 | 3592755 | 12301.58 | 49863.35 | 0.78 | 0.34 | 1.39 |
| Angola | 2012 | 36307.41 | 3628309 | 15987.87 | 64423.85 | 1.00 | 0.44 | 1.78 |
| Angola | 2013 | 40746.84 | 3856660 | 17874.42 | 72606.5 | 1.06 | 0.46 | 1.88 |
| Angola | 2014 | 43376.79 | 4180422 | 18977.15 | 77498.67 | 1.04 | 0.45 | 1.85 |
| Angola | 2015 | 46283.3 | 4603846 | 19273.58 | 84132.84 | 1.01 | 0.42 | 1.83 |
| Angola | 2016 | 56177.06 | 5131705 | 21787.16 | 103906.8 | 1.09 | 0.42 | 2.02 |
| Angola | 2017 | 74741.9 | 6020189 | 32120.61 | 134582.1 | 1.24 | 0.53 | 2.24 |
| Angola | 2018 | 80331.83 | 6701771 | 32924.64 | 146595.3 | 1.20 | 0.49 | 2.19 |
| Angola | 2019 | 93558.31 | 7220990 | 38786.27 | 169843 | 1.30 | 0.54 | 2.35 |
| Angola | 2020 | 99786.04 | 7340921 | 40406.98 | 182580.4 | 1.36 | 0.55 | 2.49 |
| Angola | 2021 | 113274.7 | 7559447 | 46116.76 | 206906.2 | 1.50 | 0.61 | 2.74 |
| Angola | 2022 | 95907.06 | 7769644 | 39254.2 | 175090.6 | 1.23 | 0.51 | 2.25 |
| Benin | 2011 | 13101.28 | 3751022 | -72358.4 | 94207.6 | 0.35 | -1.93 | 2.51 |
| Benin | 2012 | 7997.446 | 3693580 | -73072.5 | 83502.1 | 0.22 | -1.98 | 2.26 |
| Benin | 2013 | 15089.07 | 3766808 | -68374.3 | 94899.87 | 0.40 | -1.82 | 2.52 |
| Benin | 2014 | 12183.6 | 3954460 | -77980.7 | 98083.22 | 0.31 | -1.97 | 2.48 |
| Benin | 2015 | 16406.46 | 4364576 | -74020.1 | 102923.2 | 0.38 | -1.70 | 2.36 |
| Benin | 2016 | 14180.51 | 4848499 | -109269 | 129463.6 | 0.29 | -2.25 | 2.67 |
| Benin | 2017 | 19163.45 | 4980431 | -102498 | 134252.3 | 0.38 | -2.06 | 2.70 |
| Benin | 2018 | 13075.35 | 4972593 | -105968 | 124043.3 | 0.26 | -2.13 | 2.49 |
| Benin | 2019 | 11782.52 | 5033453 | -127017 | 138176.3 | 0.23 | -2.52 | 2.75 |
| Benin | 2020 | 20186.4 | 4925767 | -100857 | 134045.2 | 0.41 | -2.05 | 2.72 |
| Benin | 2021 | 18377.13 | 4985810 | -114052 | 144046.9 | 0.37 | -2.29 | 2.89 |
| Benin | 2022 | 16817.42 | 5138370 | -104147 | 132000.6 | 0.33 | -2.03 | 2.57 |
| Burkina Faso | 2011 | 248313.8 | 9681549 | 26236.03 | 475859.2 | 2.56 | 0.27 | 4.92 |
| Burkina Faso | 2012 | 213341.3 | 9584774 | 10523.85 | 423564.6 | 2.23 | 0.11 | 4.42 |
| Burkina Faso | 2013 | 227174 | 9133534 | 23777.13 | 437507.2 | 2.49 | 0.26 | 4.79 |
| Burkina Faso | 2014 | 206093.2 | 8695611 | 21917.25 | 396288.2 | 2.37 | 0.25 | 4.56 |
| Burkina Faso | 2015 | 191620.8 | 8298701 | 20775.85 | 367280.3 | 2.31 | 0.25 | 4.43 |
| Burkina Faso | 2016 | 187157.3 | 7561786 | 19936.37 | 359786.6 | 2.48 | 0.26 | 4.76 |
| Burkina Faso | 2017 | 175942 | 7259133 | 18617.58 | 337894.2 | 2.42 | 0.26 | 4.65 |
| Burkina Faso | 2018 | 169804.7 | 7103831 | 10066.69 | 333972.1 | 2.39 | 0.14 | 4.70 |
| Burkina Faso | 2019 | 178450.8 | 7069910 | 17044.84 | 345739.7 | 2.52 | 0.24 | 4.89 |
| Burkina Faso | 2020 | 175636.2 | 7188634 | 18025.06 | 338879.9 | 2.44 | 0.25 | 4.71 |
| Burkina Faso | 2021 | 177497.4 | 7148152 | 13500.28 | 345628.5 | 2.48 | 0.19 | 4.84 |
| Burkina Faso | 2022 | 172880.6 | 7328937 | 14958.38 | 335654.8 | 2.36 | 0.20 | 4.58 |
| Burundi | 2011 | -4356.28 | 1518907 | -7487.91 | -1696.13 | -0.29 | -0.49 | -0.11 |
| Burundi | 2012 | -5733.63 | 1565977 | -9928.81 | -1959.37 | -0.37 | -0.63 | -0.13 |
| Burundi | 2013 | -7599.22 | 1689022 | -13128 | -2618.36 | -0.45 | -0.78 | -0.16 |
| Burundi | 2014 | -6669.49 | 1808588 | -11543.1 | -2274.43 | -0.37 | -0.64 | -0.13 |
| Burundi | 2015 | -12045 | 2050601 | -21102.5 | -3358.12 | -0.59 | -1.03 | -0.16 |
| Burundi | 2016 | -18251.6 | 2475229 | -32300.6 | -4662.73 | -0.74 | -1.30 | -0.19 |
| Burundi | 2017 | -15524.4 | 2775617 | -26786.1 | -5435.5 | -0.56 | -0.97 | -0.20 |
| Burundi | 2018 | -17062.2 | 3273752 | -29416.8 | -6080.57 | -0.52 | -0.90 | -0.19 |
| Burundi | 2019 | -14142 | 3446000 | -24383.5 | -5428.23 | -0.41 | -0.71 | -0.16 |
| Burundi | 2020 | -22001.2 | 3689098 | -37758.5 | -8470.81 | -0.60 | -1.02 | -0.23 |
| Burundi | 2021 | -19027.7 | 3686472 | -32624.1 | -7324.32 | -0.52 | -0.88 | -0.20 |
| Burundi | 2022 | -15174.1 | 3575939 | -26034.1 | -5890.09 | -0.42 | -0.73 | -0.16 |
| Cameroon | 2011 | -3752.77 | 6203771 | -33698.5 | 25833.28 | -0.06 | -0.54 | 0.42 |
| Cameroon | 2012 | -3105.69 | 6158834 | -28880.6 | 22493.6 | -0.05 | -0.47 | 0.37 |
| Cameroon | 2013 | -4484.92 | 6542668 | -35786.6 | 25920.4 | -0.07 | -0.55 | 0.40 |
| Cameroon | 2014 | -4415.14 | 6799625 | -36236.1 | 26591.69 | -0.06 | -0.53 | 0.39 |
| Cameroon | 2015 | -2281.12 | 7005129 | -31595.5 | 26973.55 | -0.03 | -0.45 | 0.39 |
| Cameroon | 2016 | -8581.16 | 7155083 | -48179.4 | 27399.78 | -0.12 | -0.67 | 0.38 |
| Cameroon | 2017 | -6157.64 | 7021329 | -43906.2 | 30200.83 | -0.09 | -0.63 | 0.43 |
| Cameroon | 2018 | -4989.29 | 6840353 | -40779.8 | 29828.61 | -0.07 | -0.60 | 0.44 |
| Cameroon | 2019 | -7292.18 | 6812332 | -49617.3 | 33030.7 | -0.11 | -0.73 | 0.48 |
| Cameroon | 2020 | -3210.22 | 7130713 | -46577.8 | 40091.11 | -0.05 | -0.65 | 0.56 |
| Cameroon | 2021 | -7243.67 | 7795674 | -58893 | 42938.79 | -0.09 | -0.76 | 0.55 |
| Cameroon | 2022 | -5539.57 | 8202394 | -51023.4 | 39528.97 | -0.07 | -0.62 | 0.48 |
| Chad | 2011 | -37491.8 | 2854256 | -105067 | 38038.33 | -1.31 | -3.68 | 1.33 |
| Chad | 2012 | -35070.8 | 2812988 | -98074 | 35405.4 | -1.25 | -3.49 | 1.26 |
| Chad | 2013 | -40292.6 | 2857610 | -112883 | 40547.92 | -1.41 | -3.95 | 1.42 |
| Chad | 2014 | -43101.7 | 3017180 | -121020 | 43745.52 | -1.43 | -4.01 | 1.45 |
| Chad | 2015 | -44032.6 | 3125975 | -123629 | 44714.91 | -1.41 | -3.95 | 1.43 |
| Chad | 2016 | -47249.3 | 3231345 | -132547 | 47664.4 | -1.46 | -4.10 | 1.48 |
| Chad | 2017 | -47275.5 | 3376459 | -132696 | 47978.44 | -1.40 | -3.93 | 1.42 |
| Chad | 2018 | -52536.7 | 3635680 | -147487 | 53247.35 | -1.45 | -4.06 | 1.46 |
| Chad | 2019 | -52611.3 | 3652896 | -147685 | 53251.59 | -1.44 | -4.04 | 1.46 |
| Chad | 2020 | -52007.1 | 3682988 | -145966 | 52752.94 | -1.41 | -3.96 | 1.43 |
| Chad | 2021 | -54445.6 | 3719523 | -152927 | 55215.71 | -1.46 | -4.11 | 1.48 |
| Chad | 2022 | -53089.1 | 3963609 | -148809 | 53784.66 | -1.34 | -3.75 | 1.36 |
| Democratic Republic of the Congo | 2011 | 75646.68 | 26121021 | -151530 | 278166.3 | 0.29 | -0.58 | 1.06 |
| Democratic Republic of the Congo | 2012 | 66557.02 | 24885064 | -127713 | 240160.3 | 0.27 | -0.51 | 0.97 |
| Democratic Republic of the Congo | 2013 | 83172.34 | 23614999 | -155531 | 292586.4 | 0.35 | -0.66 | 1.24 |
| Democratic Republic of the Congo | 2014 | 75696.01 | 23239029 | -151593 | 278143.8 | 0.33 | -0.65 | 1.20 |
| Democratic Republic of the Congo | 2015 | 86722.48 | 24154179 | -171798 | 317196 | 0.36 | -0.71 | 1.31 |
| Democratic Republic of the Congo | 2016 | 105315.1 | 25387593 | -225920 | 400908.2 | 0.41 | -0.89 | 1.58 |
| Democratic Republic of the Congo | 2017 | 111805.6 | 26948571 | -231375 | 415270.1 | 0.41 | -0.86 | 1.54 |
| Democratic Republic of the Congo | 2018 | 111286.1 | 28826414 | -212516 | 398169.7 | 0.39 | -0.74 | 1.38 |
| Democratic Republic of the Congo | 2019 | 115410.6 | 29697644 | -253091 | 443647.9 | 0.39 | -0.85 | 1.49 |
| Democratic Republic of the Congo | 2020 | 109672 | 28756401 | -273431 | 455353.3 | 0.38 | -0.95 | 1.58 |
| Democratic Republic of the Congo | 2021 | 111967.4 | 28614220 | -254486 | 438050 | 0.39 | -0.89 | 1.53 |
| Democratic Republic of the Congo | 2022 | 78345.24 | 28890697 | -209801 | 342147.4 | 0.27 | -0.73 | 1.18 |
| Ivory Coast | 2011 | 12638.71 | 10359501 | -98960.2 | 125932.3 | 0.12 | -0.96 | 1.22 |
| Ivory Coast | 2012 | 9058.593 | 8616209 | -70677.3 | 90688.67 | 0.11 | -0.82 | 1.05 |
| Ivory Coast | 2013 | 8424.551 | 6792213 | -66707.4 | 84225.32 | 0.12 | -0.98 | 1.24 |
| Ivory Coast | 2014 | 8560.864 | 6237956 | -67654.6 | 84283.49 | 0.14 | -1.08 | 1.35 |
| Ivory Coast | 2015 | 7271.613 | 6043583 | -57572.1 | 71779.54 | 0.12 | -0.95 | 1.19 |
| Ivory Coast | 2016 | 9415.902 | 6210859 | -74377.8 | 92564.3 | 0.15 | -1.20 | 1.49 |
| Ivory Coast | 2017 | 10243.82 | 6581187 | -80312.3 | 101857.3 | 0.16 | -1.22 | 1.55 |
| Ivory Coast | 2018 | 10896.53 | 7081304 | -85754.8 | 107567.8 | 0.15 | -1.21 | 1.52 |
| Ivory Coast | 2019 | 13282.88 | 7460630 | -105230 | 130362.7 | 0.18 | -1.41 | 1.75 |
| Ivory Coast | 2020 | 11884.49 | 7727820 | -94440.7 | 117030.9 | 0.15 | -1.22 | 1.51 |
| Ivory Coast | 2021 | 13855.77 | 7974179 | -109420 | 136496.8 | 0.17 | -1.37 | 1.71 |
| Ivory Coast | 2022 | 12024.56 | 8211654 | -94914.7 | 118153.8 | 0.15 | -1.16 | 1.44 |
| Gabon | 2011 | -517.131 | 337394.9 | -3046.23 | 2058.253 | -0.15 | -0.90 | 0.61 |
| Gabon | 2012 | -469.7 | 357795.6 | -2725.58 | 1802.142 | -0.13 | -0.76 | 0.50 |
| Gabon | 2013 | -618.669 | 376414 | -3596.81 | 2378.374 | -0.16 | -0.96 | 0.63 |
| Gabon | 2014 | -636.085 | 378557.8 | -3694.12 | 2431.003 | -0.17 | -0.98 | 0.64 |
| Gabon | 2015 | -526.39 | 367989.5 | -3095.49 | 2075.57 | -0.14 | -0.84 | 0.56 |
| Gabon | 2016 | -711.436 | 361655.6 | -4167.69 | 2821.248 | -0.20 | -1.15 | 0.78 |
| Gabon | 2017 | -635.319 | 361255.8 | -3719.58 | 2519.206 | -0.18 | -1.03 | 0.70 |
| Gabon | 2018 | -623.159 | 361435.6 | -3606.15 | 2383.719 | -0.17 | -1.00 | 0.66 |
| Gabon | 2019 | -685.357 | 371518.7 | -3967.7 | 2616.736 | -0.18 | -1.07 | 0.70 |
| Gabon | 2020 | -730.363 | 382189.5 | -4225.48 | 2793.494 | -0.19 | -1.11 | 0.73 |
| Gabon | 2021 | -784.197 | 381792.3 | -4528.92 | 3027.935 | -0.21 | -1.19 | 0.79 |
| Gabon | 2022 | -636.67 | 373738.5 | -3678.3 | 2441.829 | -0.17 | -0.98 | 0.65 |
| Gambia | 2011 | 18405.12 | 830014.3 | -4015.29 | 41758.44 | 2.22 | -0.48 | 5.03 |
| Gambia | 2012 | 19671.31 | 915220 | -3732.51 | 44593.02 | 2.15 | -0.41 | 4.87 |
| Gambia | 2013 | 22159.6 | 804950.2 | -3975.86 | 49450.16 | 2.75 | -0.49 | 6.14 |
| Gambia | 2014 | 12455.45 | 508641.7 | -2821.99 | 28313.05 | 2.45 | -0.55 | 5.57 |
| Gambia | 2015 | 15905.21 | 692909.7 | -2562.24 | 36096.35 | 2.30 | -0.37 | 5.21 |
| Gambia | 2016 | 10182.31 | 421254.4 | -2628.41 | 23275.44 | 2.42 | -0.62 | 5.53 |
| Gambia | 2017 | 5406.399 | 221681.3 | -1194.39 | 12257.71 | 2.44 | -0.54 | 5.53 |
| Gambia | 2018 | 5243.217 | 247677.2 | -1281.5 | 12134.26 | 2.12 | -0.52 | 4.90 |
| Gambia | 2019 | 3907.656 | 163107.4 | -957.792 | 9029.978 | 2.40 | -0.59 | 5.54 |
| Gambia | 2020 | 5840.544 | 210850.9 | -1073.58 | 13156.37 | 2.77 | -0.51 | 6.24 |
| Gambia | 2021 | 4965.467 | 204817 | -1262.86 | 11416.14 | 2.42 | -0.62 | 5.57 |
| Gambia | 2022 | 6974.251 | 309901.5 | -1565.73 | 15857.46 | 2.25 | -0.51 | 5.12 |
| Ghana | 2011 | 239945.3 | 9741136 | 141236.4 | 335814.9 | 2.46 | 1.45 | 3.45 |
| Ghana | 2012 | 220867.8 | 9697369 | 128921.2 | 310189.1 | 2.28 | 1.33 | 3.20 |
| Ghana | 2013 | 256932.5 | 9278858 | 158070.7 | 352579.7 | 2.77 | 1.70 | 3.80 |
| Ghana | 2014 | 227660.9 | 8637491 | 131358.4 | 320987.9 | 2.64 | 1.52 | 3.72 |
| Ghana | 2015 | 194196.4 | 7785362 | 115550.6 | 269458.9 | 2.49 | 1.48 | 3.46 |
| Ghana | 2016 | 194002.8 | 6750441 | 110003.6 | 275099.1 | 2.87 | 1.63 | 4.08 |
| Ghana | 2017 | 160311.2 | 5851922 | 89486.05 | 229258 | 2.74 | 1.53 | 3.92 |
| Ghana | 2018 | 144214.4 | 5199190 | 82183.35 | 204159.3 | 2.77 | 1.58 | 3.93 |
| Ghana | 2019 | 157703.6 | 5116429 | 85153.39 | 228128.1 | 3.08 | 1.66 | 4.46 |
| Ghana | 2020 | 168751.5 | 5261296 | 100520.8 | 234103 | 3.21 | 1.91 | 4.45 |
| Ghana | 2021 | 169042.2 | 5178838 | 95348.89 | 239989.3 | 3.26 | 1.84 | 4.63 |
| Ghana | 2022 | 159619.4 | 5335832 | 94666.14 | 220650.5 | 2.99 | 1.77 | 4.14 |
| Guinea | 2011 | 5055.173 | 4396820 | -24326 | 41771.13 | 0.11 | -0.55 | 0.95 |
| Guinea | 2012 | 6067.608 | 4550355 | -20862.6 | 39992.51 | 0.13 | -0.46 | 0.88 |
| Guinea | 2013 | 7437.36 | 4477373 | -28380.8 | 52490.15 | 0.17 | -0.63 | 1.17 |
| Guinea | 2014 | 10336.72 | 4412137 | -20083.7 | 50425.9 | 0.23 | -0.46 | 1.14 |
| Guinea | 2015 | 5117.325 | 4396517 | -24038 | 41409.92 | 0.12 | -0.55 | 0.94 |
| Guinea | 2016 | 6609.974 | 4314363 | -30308.1 | 52536.04 | 0.15 | -0.70 | 1.22 |
| Guinea | 2017 | 5308.912 | 4337021 | -26854.5 | 45646.71 | 0.12 | -0.62 | 1.05 |
| Guinea | 2018 | 9026.846 | 4330178 | -26357.8 | 54397.62 | 0.21 | -0.61 | 1.26 |
| Guinea | 2019 | 9098.645 | 4080472 | -28299.1 | 56449 | 0.22 | -0.69 | 1.38 |
| Guinea | 2020 | 6312.577 | 3841814 | -27735.9 | 48611.1 | 0.16 | -0.72 | 1.27 |
| Guinea | 2021 | 3806.247 | 3646688 | -27920.8 | 42818.33 | 0.10 | -0.77 | 1.17 |
| Guinea | 2022 | 4367.616 | 3635356 | -25570 | 41759.26 | 0.12 | -0.70 | 1.15 |
| Kenya | 2011 | 49646.74 | 2906411 | 35753.56 | 62723.54 | 1.71 | 1.23 | 2.16 |
| Kenya | 2012 | 47368.13 | 3109761 | 33037.89 | 61215.74 | 1.52 | 1.06 | 1.97 |
| Kenya | 2013 | 38966.34 | 3233526 | 28087.28 | 49592.86 | 1.21 | 0.87 | 1.53 |
| Kenya | 2014 | 46646.7 | 3305553 | 33627.91 | 59165.07 | 1.41 | 1.02 | 1.79 |
| Kenya | 2015 | 63482.09 | 3283376 | 45697.44 | 80262.83 | 1.93 | 1.39 | 2.44 |
| Kenya | 2016 | 78254.26 | 3133027 | 56868.77 | 97594.45 | 2.50 | 1.82 | 3.12 |
| Kenya | 2017 | 83401.16 | 3126599 | 60980.32 | 103747.5 | 2.67 | 1.95 | 3.32 |
| Kenya | 2018 | 68223.11 | 3084134 | 50132.66 | 84464.95 | 2.21 | 1.63 | 2.74 |
| Kenya | 2019 | 69505.01 | 3044510 | 50467.66 | 86888.41 | 2.28 | 1.66 | 2.85 |
| Kenya | 2020 | 56543.94 | 3274143 | 41956.58 | 69546.87 | 1.73 | 1.28 | 2.12 |
| Kenya | 2021 | 68278.3 | 3417878 | 49944.19 | 84820.53 | 2.00 | 1.46 | 2.48 |
| Kenya | 2022 | 71393.22 | 3569500 | 52178.09 | 88647.22 | 2.00 | 1.46 | 2.48 |
| Liberia | 2011 | 2749.72 | 1287757 | -6053.33 | 11933.07 | 0.21 | -0.47 | 0.93 |
| Liberia | 2012 | 1777.451 | 1241971 | -4313.99 | 8113.472 | 0.14 | -0.35 | 0.65 |
| Liberia | 2013 | 3156.013 | 1273810 | -5801.34 | 12557.41 | 0.25 | -0.46 | 0.99 |
| Liberia | 2014 | 2790.424 | 1386657 | -6988.69 | 12955.44 | 0.20 | -0.50 | 0.93 |
| Liberia | 2015 | 2390.436 | 1456101 | -7607.55 | 12943.74 | 0.16 | -0.52 | 0.89 |
| Liberia | 2016 | 2704.31 | 1587037 | -10735.9 | 16834.52 | 0.17 | -0.68 | 1.06 |
| Liberia | 2017 | 3037.362 | 1684596 | -11443.5 | 18244.57 | 0.18 | -0.68 | 1.08 |
| Liberia | 2018 | 1221.795 | 1672356 | -14635.2 | 17388.56 | 0.07 | -0.88 | 1.04 |
| Liberia | 2019 | 1187.758 | 1637674 | -19709.9 | 22553.95 | 0.07 | -1.20 | 1.38 |
| Liberia | 2020 | 2192.29 | 1630681 | -14874.7 | 19870.48 | 0.13 | -0.91 | 1.22 |
| Liberia | 2021 | 2337.576 | 1574262 | -15863.9 | 21112.13 | 0.15 | -1.01 | 1.34 |
| Liberia | 2022 | 3452.379 | 1565342 | -11779 | 19429.69 | 0.22 | -0.75 | 1.24 |
| Madagascar | 2011 | 13623.58 | 1805046 | -360.446 | 26336.57 | 0.75 | -0.02 | 1.46 |
| Madagascar | 2012 | 14875.25 | 1902515 | 485.3613 | 28024.2 | 0.78 | 0.03 | 1.47 |
| Madagascar | 2013 | 22912.59 | 2129113 | 3533.055 | 41198.41 | 1.08 | 0.17 | 1.94 |
| Madagascar | 2014 | 23156.43 | 2266177 | 1974.675 | 42866.34 | 1.02 | 0.09 | 1.89 |
| Madagascar | 2015 | 22942.55 | 2223608 | 3278.604 | 41537.97 | 1.03 | 0.15 | 1.87 |
| Madagascar | 2016 | 28969.66 | 2210769 | 6032.182 | 50860.76 | 1.31 | 0.27 | 2.30 |
| Madagascar | 2017 | 28819.39 | 2319743 | 4888.831 | 51490.15 | 1.24 | 0.21 | 2.22 |
| Madagascar | 2018 | 31065.04 | 2572139 | 7139.001 | 53985.3 | 1.21 | 0.28 | 2.10 |
| Madagascar | 2019 | 36205.09 | 3045941 | 6228.996 | 64501.93 | 1.19 | 0.20 | 2.12 |
| Madagascar | 2020 | 44251.84 | 3463828 | 10758.51 | 75195.46 | 1.28 | 0.31 | 2.17 |
| Madagascar | 2021 | 53142.65 | 3816911 | 12961.26 | 90740.31 | 1.39 | 0.34 | 2.38 |
| Madagascar | 2022 | 34805.34 | 3534042 | 6776.125 | 61232.76 | 0.98 | 0.19 | 1.73 |
| Malawi | 2011 | -47654.8 | 5295319 | -85237 | -12523.3 | -0.90 | -1.61 | -0.24 |
| Malawi | 2012 | -43371.4 | 4820505 | -77566.5 | -11390.7 | -0.90 | -1.61 | -0.24 |
| Malawi | 2013 | -41439.9 | 4458948 | -74105.1 | -9744.81 | -0.93 | -1.66 | -0.22 |
| Malawi | 2014 | -37426.7 | 4160256 | -66914.3 | -8795.08 | -0.90 | -1.61 | -0.21 |
| Malawi | 2015 | -36386.7 | 3919491 | -65134.6 | -9581.88 | -0.93 | -1.66 | -0.24 |
| Malawi | 2016 | -44168.1 | 3743159 | -78626.4 | -12328.3 | -1.18 | -2.10 | -0.33 |
| Malawi | 2017 | -34480.7 | 3736399 | -61422.4 | -9380.31 | -0.92 | -1.64 | -0.25 |
| Malawi | 2018 | -36795.3 | 3665000 | -65854 | -9654.48 | -1.00 | -1.80 | -0.26 |
| Malawi | 2019 | -35154.4 | 3675482 | -62314 | -9815.86 | -0.96 | -1.70 | -0.27 |
| Malawi | 2020 | -38593.8 | 4098587 | -68630.1 | -10629.9 | -0.94 | -1.67 | -0.26 |
| Malawi | 2021 | -40667.1 | 4153218 | -71963.3 | -11354.4 | -0.98 | -1.73 | -0.27 |
| Malawi | 2022 | -36647 | 4488429 | -65423.6 | -9654.53 | -0.82 | -1.46 | -0.22 |
| Mali | 2011 | 192848.3 | 6570249 | 5402.073 | 401039.6 | 2.94 | 0.08 | 6.10 |
| Mali | 2012 | 193419.8 | 7358978 | -19988.8 | 434196 | 2.63 | -0.27 | 5.90 |
| Mali | 2013 | 234158.6 | 7887100 | -2476.5 | 495868 | 2.97 | -0.03 | 6.29 |
| Mali | 2014 | 216811 | 7712484 | -23.9053 | 457378.6 | 2.81 | 0.00 | 5.93 |
| Mali | 2015 | 170371.5 | 6623932 | -3939.02 | 364361.8 | 2.57 | -0.06 | 5.50 |
| Mali | 2016 | 152861.7 | 5279971 | -556.996 | 322842.2 | 2.90 | -0.01 | 6.11 |
| Mali | 2017 | 133878.5 | 4636588 | -19.7189 | 282289.2 | 2.89 | 0.00 | 6.09 |
| Mali | 2018 | 128651.4 | 4450321 | -8512.61 | 281084.2 | 2.89 | -0.19 | 6.32 |
| Mali | 2019 | 142628.7 | 4771256 | 2916.203 | 297507.7 | 2.99 | 0.06 | 6.24 |
| Mali | 2020 | 138859 | 4861634 | -3565.1 | 296396.7 | 2.86 | -0.07 | 6.10 |
| Mali | 2021 | 150044.8 | 4870423 | -4166.17 | 319829.1 | 3.08 | -0.09 | 6.57 |
| Mali | 2022 | 164758.5 | 5730411 | -8525.8 | 355974.8 | 2.88 | -0.15 | 6.21 |
| Mozambique | 2011 | -36618.8 | 9094965 | -82947.7 | 8772.751 | -0.40 | -0.91 | 0.10 |
| Mozambique | 2012 | -37546.1 | 9248332 | -85813.2 | 10276.85 | -0.41 | -0.93 | 0.11 |
| Mozambique | 2013 | -36466.4 | 9437429 | -84349 | 10333.23 | -0.39 | -0.89 | 0.11 |
| Mozambique | 2014 | -37094.3 | 9471171 | -84848.2 | 10193.3 | -0.39 | -0.90 | 0.11 |
| Mozambique | 2015 | -39222.1 | 9446301 | -89097 | 10421.79 | -0.42 | -0.94 | 0.11 |
| Mozambique | 2016 | -45836.1 | 9452421 | -103838 | 10828.52 | -0.48 | -1.10 | 0.11 |
| Mozambique | 2017 | -39348.5 | 9472960 | -89373.4 | 10414.47 | -0.42 | -0.94 | 0.11 |
| Mozambique | 2018 | -37591.5 | 9484967 | -86155 | 10460.67 | -0.40 | -0.91 | 0.11 |
| Mozambique | 2019 | -41731.4 | 9468555 | -92987.1 | 8787.971 | -0.44 | -0.98 | 0.09 |
| Mozambique | 2020 | -43068.6 | 9922842 | -97747.1 | 11049.86 | -0.43 | -0.99 | 0.11 |
| Mozambique | 2021 | -42203.6 | 9921348 | -96272 | 11450.43 | -0.43 | -0.97 | 0.12 |
| Mozambique | 2022 | -40396.2 | 10252885 | -91505 | 9702.974 | -0.39 | -0.89 | 0.09 |
| Niger | 2011 | 27511.95 | 6334426 | -137578 | 246158.9 | 0.43 | -2.17 | 3.89 |
| Niger | 2012 | 23821.75 | 6500223 | -136063 | 237853.4 | 0.37 | -2.09 | 3.66 |
| Niger | 2013 | 26819.65 | 6687772 | -146620 | 260529.8 | 0.40 | -2.19 | 3.90 |
| Niger | 2014 | 31594.52 | 6840288 | -155351 | 278826.4 | 0.46 | -2.27 | 4.08 |
| Niger | 2015 | 24948.6 | 6867034 | -137973 | 245139.1 | 0.36 | -2.01 | 3.57 |
| Niger | 2016 | 19906.81 | 6740268 | -147831 | 249215.4 | 0.30 | -2.19 | 3.70 |
| Niger | 2017 | 25609.17 | 6824219 | -139901 | 249307.8 | 0.38 | -2.05 | 3.65 |
| Niger | 2018 | 23256.29 | 6983167 | -148439 | 254353.8 | 0.33 | -2.13 | 3.64 |
| Niger | 2019 | 29245.59 | 7297866 | -167486 | 291572.9 | 0.40 | -2.29 | 4.00 |
| Niger | 2020 | 27019.6 | 7567702 | -163841 | 283829.7 | 0.36 | -2.17 | 3.75 |
| Niger | 2021 | 35777.83 | 7820534 | -177710 | 317142.2 | 0.46 | -2.27 | 4.06 |
| Niger | 2022 | 24826.39 | 6743862 | -143024 | 249671.1 | 0.37 | -2.12 | 3.70 |
| Nigeria | 2011 | 998570 | 59802112 | 265229.3 | 1801299 | 1.67 | 0.44 | 3.01 |
| Nigeria | 2012 | 948990.5 | 57775657 | 257474.6 | 1685107 | 1.64 | 0.45 | 2.92 |
| Nigeria | 2013 | 1041961 | 56076010 | 282462.3 | 1848380 | 1.86 | 0.50 | 3.30 |
| Nigeria | 2014 | 1081719 | 55828666 | 298050.4 | 1913910 | 1.94 | 0.53 | 3.43 |
| Nigeria | 2015 | 965352.3 | 56122164 | 257957.6 | 1740556 | 1.72 | 0.46 | 3.10 |
| Nigeria | 2016 | 1160721 | 56386955 | 332247.3 | 2026905 | 2.06 | 0.59 | 3.59 |
| Nigeria | 2017 | 1106840 | 59026019 | 297929.8 | 1971959 | 1.88 | 0.50 | 3.34 |
| Nigeria | 2018 | 1166040 | 62281678 | 336393.8 | 2026350 | 1.87 | 0.54 | 3.25 |
| Nigeria | 2019 | 1327622 | 64574915 | 388227.1 | 2298362 | 2.06 | 0.60 | 3.56 |
| Nigeria | 2020 | 1433376 | 70249023 | 409695.1 | 2505380 | 2.04 | 0.58 | 3.57 |
| Nigeria | 2021 | 1491278 | 70907574 | 404947.8 | 2660177 | 2.10 | 0.57 | 3.75 |
| Nigeria | 2022 | 1310517 | 69808842 | 377810.7 | 2280497 | 1.88 | 0.54 | 3.27 |
| Rwanda | 2011 | -784.515 | 822314.4 | -1584.94 | 37.77943 | -0.10 | -0.19 | 0.00 |
| Rwanda | 2012 | -1251.15 | 742937.3 | -2529.43 | 55.88712 | -0.17 | -0.34 | 0.01 |
| Rwanda | 2013 | -1210.34 | 647250.4 | -2447.29 | 54.70771 | -0.19 | -0.38 | 0.01 |
| Rwanda | 2014 | -831.037 | 625145.5 | -1681.28 | 38.68931 | -0.13 | -0.27 | 0.01 |
| Rwanda | 2015 | -2540.79 | 671272.8 | -5145.79 | 111.8362 | -0.38 | -0.77 | 0.02 |
| Rwanda | 2016 | -3495.75 | 776347.8 | -7093.54 | 183.7629 | -0.45 | -0.91 | 0.02 |
| Rwanda | 2017 | -3428.41 | 890086.9 | -6955.76 | 152.2667 | -0.39 | -0.78 | 0.02 |
| Rwanda | 2018 | -2121.64 | 917244.1 | -4295.15 | 95.28521 | -0.23 | -0.47 | 0.01 |
| Rwanda | 2019 | -1757.81 | 803212.2 | -3555.78 | 79.06717 | -0.22 | -0.44 | 0.01 |
| Rwanda | 2020 | -1685.88 | 686051.7 | -3410.13 | 104.2993 | -0.25 | -0.50 | 0.02 |
| Rwanda | 2021 | -1677 | 728793.4 | -3398.63 | 93.86521 | -0.23 | -0.47 | 0.01 |
| Rwanda | 2022 | -1046.47 | 732879.4 | -2117.39 | 60.09297 | -0.14 | -0.29 | 0.01 |
| Senegal | 2011 | 36851.41 | 1401985 | -33102.5 | 102842 | 2.63 | -2.36 | 7.34 |
| Senegal | 2012 | 44378.71 | 1593266 | -32244.5 | 115251.9 | 2.79 | -2.02 | 7.23 |
| Senegal | 2013 | 41663.99 | 1349388 | -28762.4 | 105837.9 | 3.09 | -2.13 | 7.84 |
| Senegal | 2014 | 24373.97 | 879488.2 | -19458.8 | 65761.45 | 2.77 | -2.21 | 7.48 |
| Senegal | 2015 | 39267.73 | 1385465 | -26882.2 | 99754.29 | 2.83 | -1.94 | 7.20 |
| Senegal | 2016 | 25316.51 | 937133.1 | -20201.2 | 68377.08 | 2.70 | -2.16 | 7.30 |
| Senegal | 2017 | 27040.13 | 1019404 | -22682 | 74100.17 | 2.65 | -2.23 | 7.27 |
| Senegal | 2018 | 31150.58 | 1380201 | -33700.1 | 91345.47 | 2.26 | -2.44 | 6.62 |
| Senegal | 2019 | 23559.01 | 963721.1 | -25138.4 | 68750.12 | 2.44 | -2.61 | 7.13 |
| Senegal | 2020 | 32556.51 | 1229772 | -27394.4 | 89450 | 2.65 | -2.23 | 7.27 |
| Senegal | 2021 | 40814.6 | 1468877 | -37858.7 | 114711.1 | 2.78 | -2.58 | 7.81 |
| Senegal | 2022 | 27102.85 | 1059873 | -26034.5 | 77034.26 | 2.56 | -2.46 | 7.27 |
| Sierra Leone | 2011 | 29522.51 | 2690150 | 1905.512 | 56167.84 | 1.10 | 0.07 | 2.09 |
| Sierra Leone | 2012 | 18738.81 | 2696331 | 2263.377 | 34961.45 | 0.69 | 0.08 | 1.30 |
| Sierra Leone | 2013 | 41031.95 | 2658388 | 5941.01 | 75240 | 1.54 | 0.22 | 2.83 |
| Sierra Leone | 2014 | 37601.93 | 2626279 | 7164.226 | 66213.22 | 1.43 | 0.27 | 2.52 |
| Sierra Leone | 2015 | 28128.28 | 2645282 | 3559.026 | 52076.98 | 1.06 | 0.13 | 1.97 |
| Sierra Leone | 2016 | 56335.44 | 2723375 | 10691.05 | 99053.22 | 2.07 | 0.39 | 3.64 |
| Sierra Leone | 2017 | 32137.11 | 2711935 | 136.5237 | 63258.77 | 1.19 | 0.01 | 2.33 |
| Sierra Leone | 2018 | 37961.85 | 2655931 | 4905.296 | 70080.53 | 1.43 | 0.18 | 2.64 |
| Sierra Leone | 2019 | 54435.65 | 2533458 | 8021.735 | 99398.26 | 2.15 | 0.32 | 3.92 |
| Sierra Leone | 2020 | 43637.95 | 2507183 | 2302.276 | 83233.46 | 1.74 | 0.09 | 3.32 |
| Sierra Leone | 2021 | 37664.1 | 2473771 | -573.835 | 75081.72 | 1.52 | -0.02 | 3.04 |
| Sierra Leone | 2022 | 36861.12 | 2472150 | 3076.387 | 69696.93 | 1.49 | 0.12 | 2.82 |
| United Republic of Tanzania | 2011 | 118266.8 | 5887580 | 61683.5 | 170872.3 | 2.01 | 1.05 | 2.90 |
| United Republic of Tanzania | 2012 | 103178.2 | 5566742 | 54445.56 | 149527.2 | 1.85 | 0.98 | 2.69 |
| United Republic of Tanzania | 2013 | 119857.7 | 6235213 | 62491.67 | 173454.4 | 1.92 | 1.00 | 2.78 |
| United Republic of Tanzania | 2014 | 134258.1 | 7086821 | 69930.98 | 194991.7 | 1.89 | 0.99 | 2.75 |
| United Republic of Tanzania | 2015 | 138224.7 | 7126427 | 72859.07 | 200174.1 | 1.94 | 1.02 | 2.81 |
| United Republic of Tanzania | 2016 | 153763.7 | 6973081 | 81200.9 | 222175.9 | 2.21 | 1.16 | 3.19 |
| United Republic of Tanzania | 2017 | 147723.7 | 6599619 | 77994.37 | 213422.1 | 2.24 | 1.18 | 3.23 |
| United Republic of Tanzania | 2018 | 111154.3 | 6259975 | 58749.3 | 160827.4 | 1.78 | 0.94 | 2.57 |
| United Republic of Tanzania | 2019 | 126115.9 | 6685302 | 66739.06 | 182131.4 | 1.89 | 1.00 | 2.72 |
| United Republic of Tanzania | 2020 | 111674.8 | 7152496 | 57998.17 | 162521.9 | 1.56 | 0.81 | 2.27 |
| United Republic of Tanzania | 2021 | 153093.5 | 8005440 | 81005.58 | 221153.2 | 1.91 | 1.01 | 2.76 |
| United Republic of Tanzania | 2022 | 167593.8 | 8868711 | 87858.26 | 242527.3 | 1.89 | 0.99 | 2.73 |
| Togo | 2011 | 2350.488 | 2247000 | -42601.3 | 51717.34 | 0.10 | -1.90 | 2.30 |
| Togo | 2012 | -1483.49 | 2520195 | -48751.4 | 51137.61 | -0.06 | -1.93 | 2.03 |
| Togo | 2013 | 3588.148 | 2838941 | -53609.3 | 66568.73 | 0.13 | -1.89 | 2.34 |
| Togo | 2014 | 701.2352 | 2933573 | -60474.7 | 68894.81 | 0.02 | -2.06 | 2.35 |
| Togo | 2015 | 2999.933 | 2899344 | -53588.6 | 65119.11 | 0.10 | -1.85 | 2.25 |
| Togo | 2016 | 430.8981 | 2706854 | -59029.8 | 66352.93 | 0.02 | -2.18 | 2.45 |
| Togo | 2017 | -163.751 | 2371968 | -53035.7 | 57909.97 | -0.01 | -2.24 | 2.44 |
| Togo | 2018 | 237.0877 | 2177068 | -46046.1 | 51443.64 | 0.01 | -2.12 | 2.36 |
| Togo | 2019 | -678.354 | 2094964 | -51108.7 | 55215.31 | -0.03 | -2.44 | 2.64 |
| Togo | 2020 | 2294.072 | 2164497 | -47329.6 | 56969.08 | 0.11 | -2.19 | 2.63 |
| Togo | 2021 | 2263.999 | 2122400 | -50140.9 | 60071.73 | 0.11 | -2.36 | 2.83 |
| Togo | 2022 | 2169.373 | 2210968 | -46266.4 | 55636.4 | 0.10 | -2.09 | 2.52 |
| Uganda | 2011 | 14501.81 | 13361527 | -44490.7 | 73978.64 | 0.11 | -0.33 | 0.55 |
| Uganda | 2012 | 14291.2 | 12695793 | -43604.6 | 72454.76 | 0.11 | -0.34 | 0.57 |
| Uganda | 2013 | 11325.72 | 11554110 | -28763.4 | 51040.62 | 0.10 | -0.25 | 0.44 |
| Uganda | 2014 | 12471.36 | 11063512 | -37072.6 | 62253.67 | 0.11 | -0.34 | 0.56 |
| Uganda | 2015 | 13963.63 | 10332804 | -40801.1 | 69056.55 | 0.14 | -0.39 | 0.67 |
| Uganda | 2016 | 22665.86 | 11080056 | -54750.1 | 100008.4 | 0.20 | -0.49 | 0.90 |
| Uganda | 2017 | 20915.35 | 11197057 | -55415.9 | 97528.46 | 0.19 | -0.49 | 0.87 |
| Uganda | 2018 | 15791.87 | 10803574 | -41342.8 | 73213.5 | 0.15 | -0.38 | 0.68 |
| Uganda | 2019 | 24667.23 | 11264249 | -48550.4 | 96258.91 | 0.22 | -0.43 | 0.85 |
| Uganda | 2020 | 13645.55 | 11478994 | -20216.1 | 48350.63 | 0.12 | -0.18 | 0.42 |
| Uganda | 2021 | 16659.15 | 11485281 | -37687.8 | 72018.76 | 0.15 | -0.33 | 0.63 |
| Uganda | 2022 | 19205.82 | 11895683 | -37629.6 | 74833.49 | 0.16 | -0.32 | 0.63 |
| Zambia | 2011 | 21460.98 | 2572985 | 8224.015 | 37094.01 | 0.83 | 0.32 | 1.44 |
| Zambia | 2012 | 26723.6 | 2949820 | 10218.26 | 46218.3 | 0.91 | 0.35 | 1.57 |
| Zambia | 2013 | 33355.72 | 3465766 | 12749.76 | 57490.26 | 0.96 | 0.37 | 1.66 |
| Zambia | 2014 | 35013.37 | 3678064 | 13381.98 | 60398.73 | 0.95 | 0.36 | 1.64 |
| Zambia | 2015 | 32808.43 | 3596443 | 12651.38 | 57123.3 | 0.91 | 0.35 | 1.59 |
| Zambia | 2016 | 34279.59 | 3497246 | 13113.46 | 59050.55 | 0.98 | 0.37 | 1.69 |
| Zambia | 2017 | 25393.93 | 3355808 | 9498.568 | 44367.72 | 0.76 | 0.28 | 1.32 |
| Zambia | 2018 | 32248.91 | 3445357 | 11590.4 | 55827 | 0.94 | 0.34 | 1.62 |
| Zambia | 2019 | 36652.61 | 3448911 | 14012.93 | 63159.16 | 1.06 | 0.41 | 1.83 |
| Zambia | 2020 | 32737.26 | 3504285 | 12479.25 | 57087.15 | 0.93 | 0.36 | 1.63 |
| Zambia | 2021 | 37431.24 | 3578136 | 14285.94 | 65114.13 | 1.05 | 0.40 | 1.82 |
| Zambia | 2022 | 30773.66 | 3742399 | 11608.16 | 53786.63 | 0.82 | 0.31 | 1.44 |
| Zimbabwe | 2011 | -813.213 | 1376020 | -11536.2 | 9580.577 | -0.06 | -0.84 | 0.70 |
| Zimbabwe | 2012 | -829.122 | 1226519 | -12042.2 | 9995.213 | -0.07 | -0.98 | 0.81 |
| Zimbabwe | 2013 | -1133.17 | 1785993 | -16404.4 | 13630.14 | -0.06 | -0.92 | 0.76 |
| Zimbabwe | 2014 | -1369.15 | 2232783 | -18716.5 | 15445.72 | -0.06 | -0.84 | 0.69 |
| Zimbabwe | 2015 | -1321.16 | 2059962 | -19952.3 | 16655.92 | -0.06 | -0.97 | 0.81 |
| Zimbabwe | 2016 | -1049.1 | 1541251 | -17152.2 | 14373.57 | -0.07 | -1.11 | 0.93 |
| Zimbabwe | 2017 | -1368.31 | 2271873 | -17204.2 | 13691.56 | -0.06 | -0.76 | 0.60 |
| Zimbabwe | 2018 | -1099.48 | 1388317 | -13378.6 | 10446.24 | -0.08 | -0.96 | 0.75 |
| Zimbabwe | 2019 | -1091.88 | 1525583 | -16523.5 | 13770.57 | -0.07 | -1.08 | 0.90 |
| Zimbabwe | 2020 | -1023.78 | 1864949 | -17000.1 | 14391.68 | -0.05 | -0.91 | 0.77 |
| Zimbabwe | 2021 | -397.026 | 886512.4 | -7229.41 | 6300.034 | -0.04 | -0.82 | 0.71 |
| Zimbabwe | 2022 | -341.433 | 829233.4 | -5776.83 | 4934.5 | -0.04 | -0.70 | 0.60 |

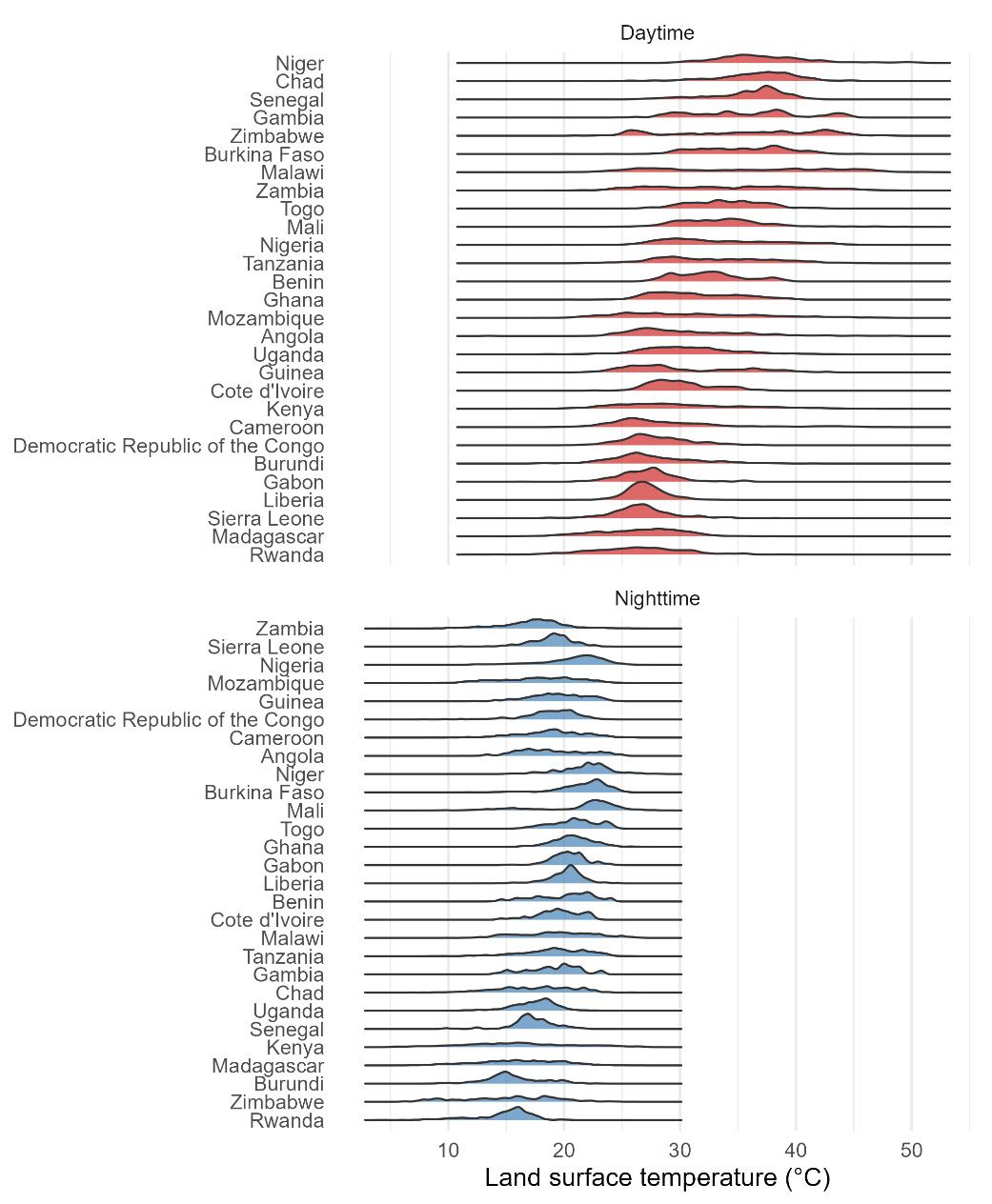

**Supplemental Figure 1.** Distributions of daytime and nighttime land surface temperatures by country.

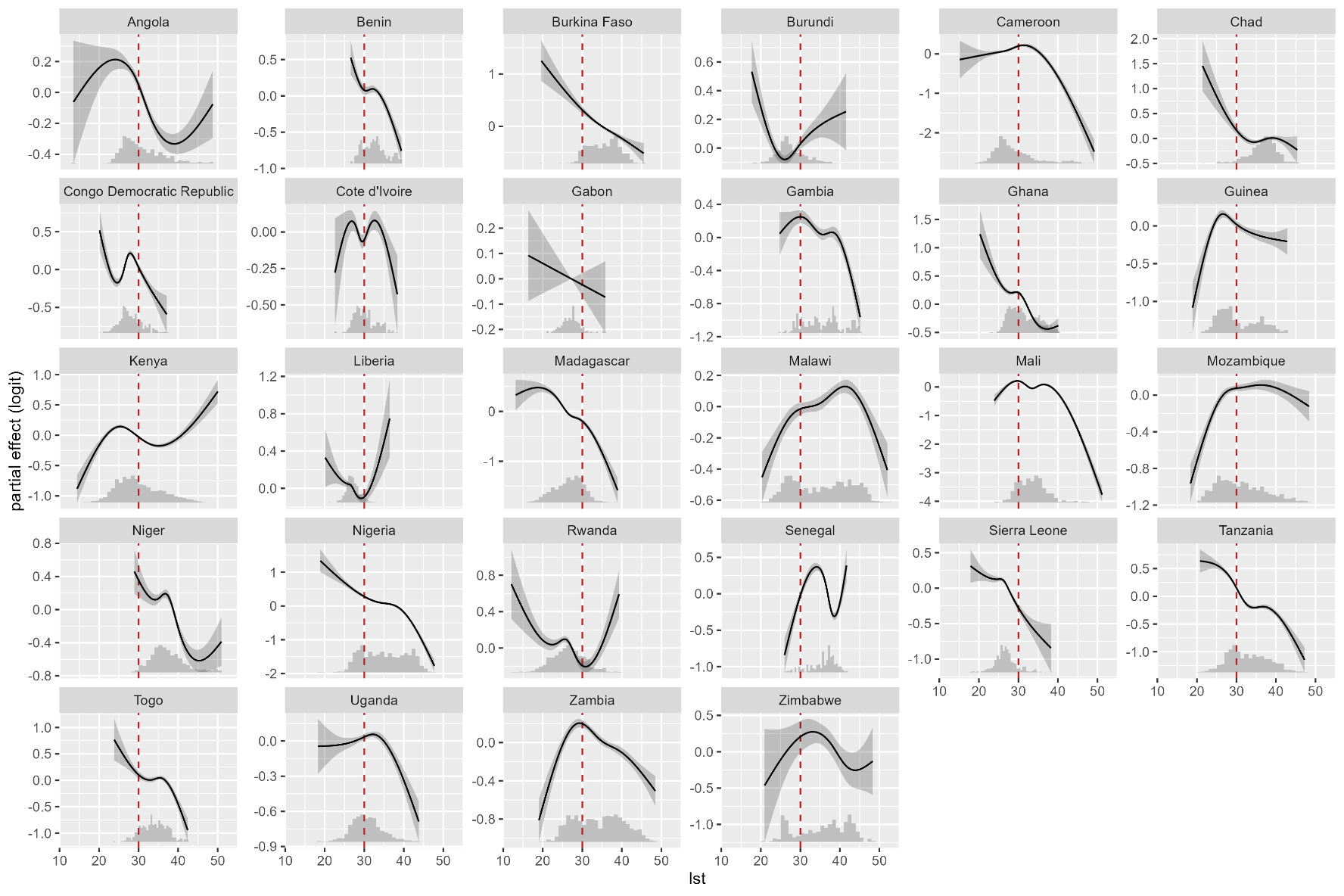

**Supplemental Figure 2.** Partial smooths resulting from country-level generalized additive models demonstrating the relationships between daytime land surface temperature and ITN use.

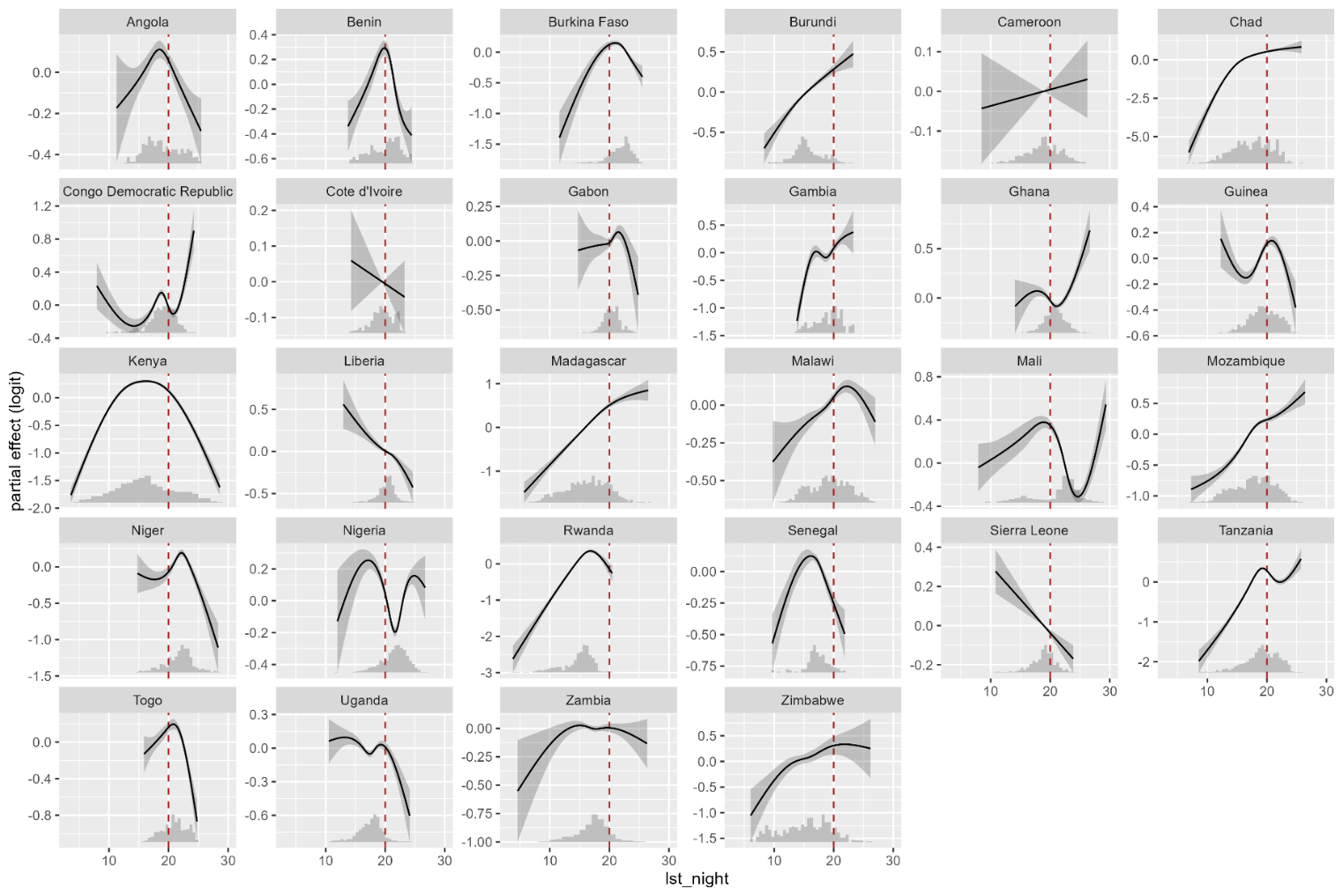

**Supplemental Figure 3.** Partial smooths resulting from country-level generalized additive models demonstrating the relationships between nighttime land surface temperature and ITN use.

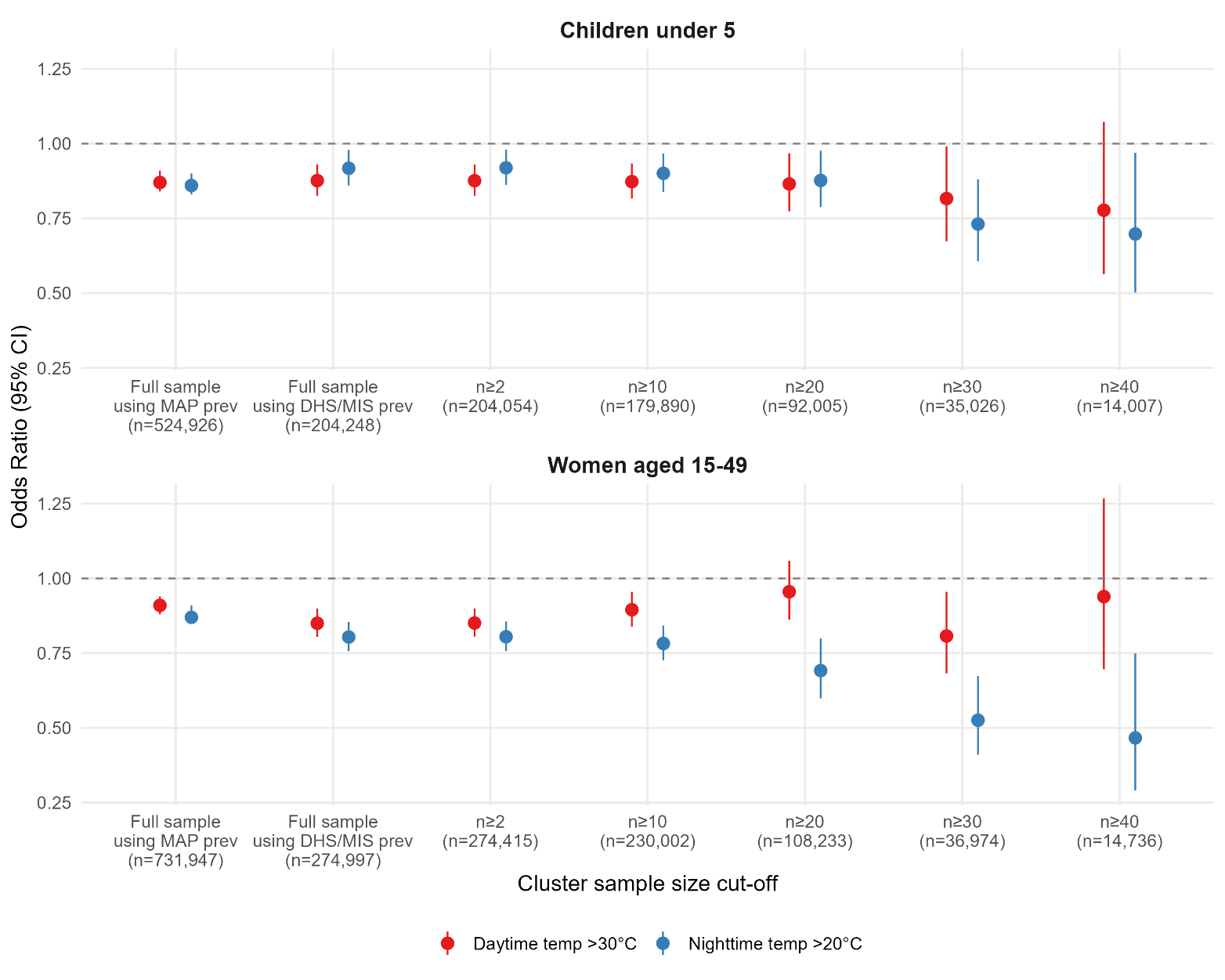

**Supplemental Figure 4.** Sensitivity analysis comparing odds ratios for ITN use at high daytime (>30°C) and nighttime (>20°C) temperatures across model specifications adjusting for cluster-level malaria prevalence, among children under 5 and women aged 15-49. The first column shows the main analysis using Malaria Atlas Project (MAP) modeled prevalence with the full sample. Subsequent columns replace MAP prevalence with cluster-level prevalence measured directly via rapid diagnostic tests or microscopy within DHS/MIS surveys, with increasingly stringent minimum cluster sample size thresholds (n≥2 to n≥40) to improve the precision of prevalence estimates. Point estimates and 95% confidence intervals are shown.
